# Aqueous Humor Liquid Biopsy Enables Multi-Omics Tumor Profiling and Methylation-Based Machine-Learning Stratification of Retinoblastoma

**DOI:** 10.64898/2026.07.09.26357661

**Authors:** Stefanie Volz, Sophia H. Montigel, Tatsiana Ryl, Elena Afanasyeva, Daniel Haag, Pia Reyes, Jan Müller, Pitithat Puranachot, Tatjana Wedig, Nathalie Schwarz, Monika Mauermann, Iman Sadeghi Dehcheshmeh, Martin Sill, Robert J. Autry, Felix Sahm, Eva Biewald, Saskia Ting, Maike Busch, Leyla Jabbarli, Tobias Kiefer, Nikolaos Bechrakis, Stefan M. Pfister, Kristian W. Pajtler, Petra Ketteler, Kendra K. Maass

## Abstract

Primary tumor biopsy in retinoblastoma carries an unacceptable risk of extraocular dissemination. As a result, children treated with eye-sparing approaches currently lack access to tumor-derived genomic information at diagnosis, limiting accurate risk stratification, preventing subtype-guided therapy, and obscuring insight into tumor evolution during conservative treatment. Aqueous humor (AH) liquid biopsy has emerged as a promising window into circulating tumor DNA (ctDNA) from eyes managed conservatively, yet its ability to comprehensively capture the genomic and epigenomic landscape of retinoblastoma and to deliver clinically actionable molecular stratification has not been rigorously evaluated. We analyzed 18 matched AH–tumor pairs using genome-wide methylation profiling, copy-number analysis, and targeted sequencing. AH samples consistently contained high ctDNA fractions (median 0.65), enabling robust detection of single-nucleotide variants, canonical copy-number alterations, and methylation signatures defining established retinoblastoma subtypes. Importantly, promoter methylation patterns associated with RB1 inactivation and optic nerve invasion were confidently detected in AH, highlighting that liquid biopsy enables functional interrogation of disease-relevant genes and pathways. To enable biopsy-independent molecular classification, we developed a methylation-based machine learning classifier trained on combined AH and tumor datasets (n=114). The classifier demonstrated exceptional performance, with AUCs of 0.96–1.00 in cross-validation and 0.97–1.00 in independent validation across 63 additional retinoblastoma cases. Together, these findings position AH liquid biopsy as powerful, minimally invasive platform for comprehensive molecular profiling in retinoblastoma. This work establishes the first clinically viable non-invasive molecular stratification tool for the disease, enabling pretreatment risk assessment and paving the way for next-generation precision diagnostics in eye-preserving care.

**One Sentence Summary:** Aqueous humor liquid biopsy enables accurate multi-omics profiling of retinoblastoma and minimally-invasive molecular risk stratification.

## INTRODUCTION

Retinoblastoma remains one of the few solid tumors for which pretreatment molecular information is essentially inaccessible. This obstructs the application of precision oncology approaches that are now routine in many other pediatric cancers. In high-resource countries, most children present with intraocular disease (*1*). Although the overall survival of children with intraocular disease is high, most are left with impaired vision or blindness and treatment related second malignancies remain a serious long-term sequela (*2*). Especially, as about 35% of children present with bilateral disease due to a genetic predisposition (*3*). Treatment options for localized intraocular retinoblastoma range from enucleation of the affected eyes to different eye-preserving modalities. However, the integration of molecular profiling into retinoblastoma research and clinical care is limited by the fundamental restriction that the tumor cannot be biopsied without risking extraocular dissemination (*4–8*). The same anatomic structures that make the eye an effective sanctuary against metastatic spread simultaneously prevent direct tumor sampling in patients undergoing eye-preserving therapies.

Consequently, the understanding of the molecular landscape of retinoblastoma has been shaped entirely by analyses of enucleated eyes (*9–13*). These specimens offer valuable insights into retinoblastoma gene (*RB1*)-driven oncogenesis, characteristic genomic and genetic alterations driving retinoblastoma tumorigenesis and subtypes but they represent more advanced disease stages and a selected patient subset.

Retinoblastoma originates in postmitotic cone photoreceptor precursor cells and manifests as unilateral or bilateral retinoblastoma (*14*). Recurrent copy number variations (CNVs) in retinoblastoma are gains on chromosomes 1q, 2p (*MYCN* locus) and 6p, and losses on 13q (*RB1* locus) and 16q (*11, 12*). Several of these alterations have been associated with aggressive clinical behavior, including an increased risk of extraocular extension, metastatic spread and resistance to chemotherapy. For instance, gain of 6p has been associated with poor ocular salvage rates, whereas *MYCN* amplification *RB1*^wt/wt^ tumors are rapidly growing, poorly differentiated tumors presenting at an earlier age (*15, 16*). Advanced multi-omics studies have further categorized retinoblastoma into at least two subtypes. Liu et al. first described two molecular subtypes based on transcriptomic, methylome and CNV differences with subtype 1 being characterized by few alterations beyond *RB1* loss and lower metastasis potential. In contrast, subtype 2 tumors exhibit more CNVs and include *MYCN*-driven tumors with stemness characteristics (*9*). Ryl et al. utilized methylation profiling and expanded the classification to three clusters: Cluster A is associated with subtype 1, while the former subtype 2 was further subdivided into Clusters B and C. Cluster C is defined by global hypomethylation compared to Cluster A and B and comprises *MYCN*-driven tumors (*10*). Unilateral sporadic retinoblastomas account for 65% of cases and are initiated by biallelic somatic inactivation of *RB1* (*17, 18*). Children with heritable retinoblastoma, that mostly presents as bilateral disease, carry a constitutional alteration of *RB1*, so that only one somatic ‘second hit’ initiates tumorigenesis. Notably, these heritable forms manifest at a younger median age of 0.46 years than non-heritable cases (median age at diagnosis 1.77 years) (*17*). In a minority of unilateral retinoblastoma *RB1* is proficient and instead, *MYCN* amplification is observed in these cases (*13*). Most children undergoing enucleation are cured (*19, 20*). While unquestionably beneficial for patients the resulting paucity of metastatic or relapsing events prevents correlation of molecular features with clinical outcomes. This limits the identification of prognostic biomarkers, undermines efforts to define molecular risk groups, and restricts the development of biologically informed therapeutic strategies. Ultimately, it is still not possible to reliably determine which patients need enucleation for effective disease control and which can be safely managed with eye-preserving therapies, nor to identify the most appropriate eye-preserving modality for an individual patient.

The shift towards eye-preserving therapies further widened this diagnostic gap (*21*). Most children treated with conservative modalities such as intra-arterial or intravitreal chemotherapy or local destructive therapies retain their eyes and do not provide tumor tissue for analysis. As a result, the biology of the majority of contemporary cases, particularly those with vision-salvageable disease, is poorly represented in current molecular datasets. The absence of molecular information also limits our understanding of tumor evolution under selective pressure from local chemotherapy and obscures mechanisms of resistance, therapy failure, and late recurrence.

Liquid biopsy offers a minimally invasive solution by detecting tumor-derived analytes **in** body fluids, addressing this longstanding limitation. Most commonly blood plasma or cerebrospinal fluid are analyzed to enable real-time therapy monitoring and to identify minimally residual disease (*22, 23*). Circulating tumor DNA (ctDNA) has emerged as the most promising analyte in liquid biopsies due to its stability and the possibility to detect somatic alterations. ctDNA represents the tumor-derived fraction within the total cell-free DNA (cfDNA) pool, the latter being released by various healthy tissues and cell populations. cfDNA is typically around 166 base pairs (bp) long, corresponding to the length of DNA wrapped around one nucleosome. In contrast, ctDNA has been found to be characteristically shorter (∼132 – 145 bp) (*24, 25*).

In retinoblastoma patients, ctDNA can be detected in aqueous humor (AH), cerebrospinal fluid and peripheral blood, enabling minimally invasive access to tumor genetics (*26–30*). Early studies demonstrate that these samples capture key driver alterations, reflect intraocular tumor burden, and may serve as real-time indicators of therapeutic response (*28, 31–34*). Importantly, liquid biopsy has the potential to profile tumors at presentation, before any treatment is administered, providing the molecular context needed to refine risk stratification and inform individualized therapeutic decisions (*27*). However, studies have been limited by small cohorts, a focus on targeted alteration detection, or have lacked integration of genetic, epigenetic, and structural alterations. To date, no study has evaluated whether AH-ctDNA can comprehensively and clinically actionably recapitulate established genomic and methylation-based retinoblastoma subtypes.

By permitting access to the molecular features of tumors undergoing eye-preserving treatments without compromising patient safety (*35*), liquid biopsy represents a transformative opportunity for retinoblastoma research and care. It may allow the field to bridge the divide between biological discovery and clinical translation, enabling the development of targeted, less toxic therapies while advancing our understanding of tumor evolution in the era of eye-sparing treatment.

The present study aims to address the critical need for minimally invasive molecular diagnostics in retinoblastoma by leveraging AH liquid biopsies. Specifically, we evaluate the feasibility of using AH-derived cfDNA for molecular tumor classification, with a focus on its ability to reflect key genomic and epigenomic tumor features. By leveraging comprehensive molecular profiling of AH cfDNA using methylation clustering, CNVs, and mutation analyses, this study seeks to establish minimally invasive diagnostic tools for retinoblastoma and pave the way for translation of liquid biopsies into the clinical management of retinoblastomas.

## RESULTS

### Patient cohort and study design

We analyzed 18 matched tumor–AH pairs from retinoblastoma patients after enucleation and subjected the resulting cfDNA to targeted panel sequencing, low-coverage whole-genome sequencing (lcWGS), methylation array and whole-genome methylation sequencing (WGMS) to derive single-nucleotide variants (SNVs), CNVs and genome-wide DNA-methylation profiles (Fig. 1A). lcWGS and methylation array analyses were performed on tumor tissue DNA to obtain CNVs and methylation patterns for direct comparison with the paired AH profiles. Extracting features derived from methylation array and sequencing including both AH and tumor profiles as well as publicly available reference data, we trained a classifier to assign samples to the retinoblastoma methylation clusters defined by Ryl et al. (*10*)(Fig. 1A).

**Figure 1.**
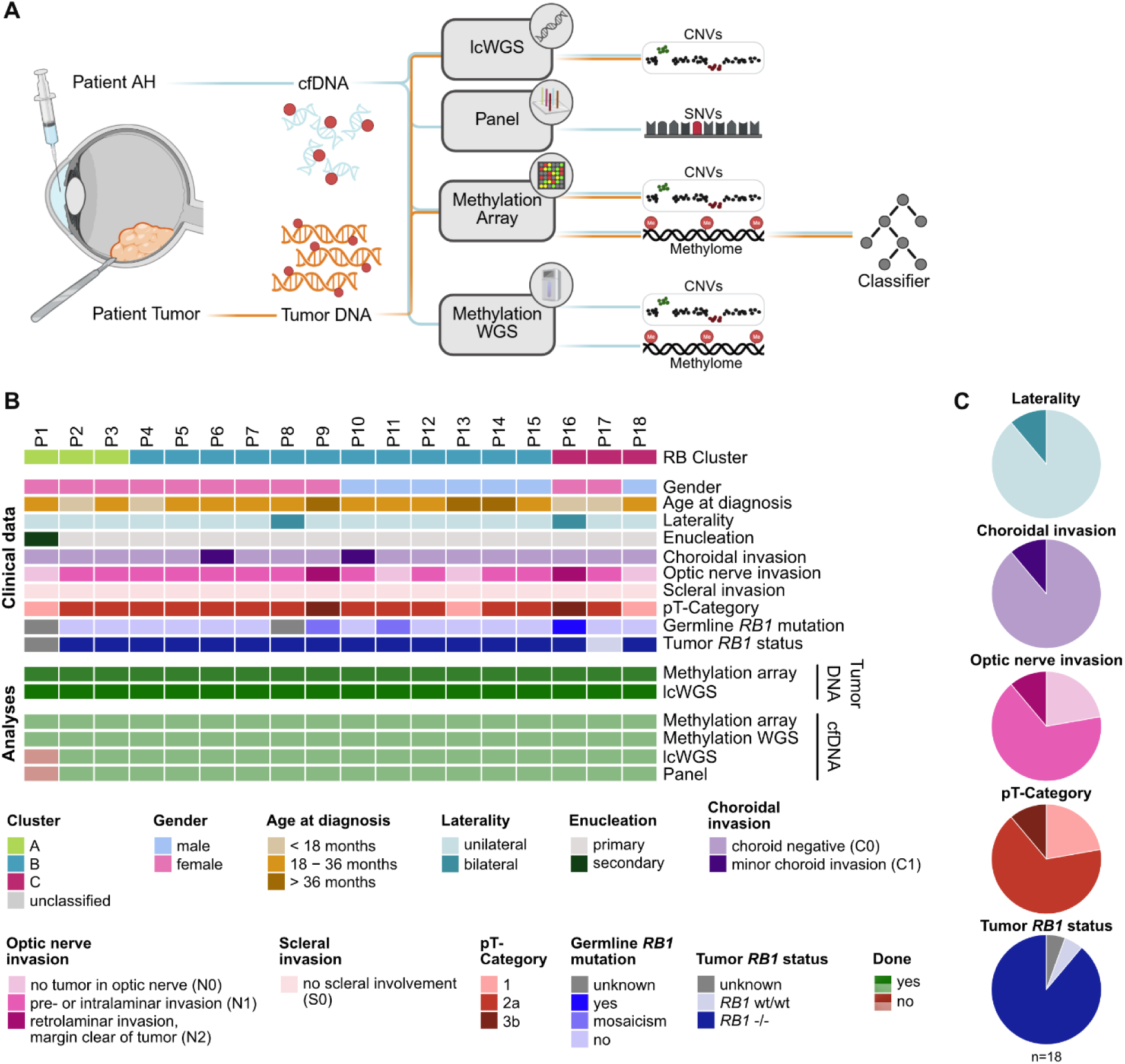
Integrated analysis of matched tumor and aqueous humor samples in retinoblastoma. **(A)** Schematic overview of the study design. Matched tumor tissue and AH samples were collected from retinoblastoma patients. cfDNA extracted from AH was subjected to lcWGS, targeted panel sequencing, methylation array, and methylation WGS. Tumor DNA underwent WGS and methylation array profiling. CNVs, SNVs, and methylation profiles were derived accordingly, and methylation-based data were used to train a classifier for tumor stratification. **(B)** Cohort overview plot summarizing the retinoblastoma cohort (n=18), annotated with retinoblastoma cluster classification, clinical features, and tumor characteristics. Molecular data availability for each patient is shown below, including analyses performed on tumor and/or AH cfDNA samples. Staging of tumors was performed according to IRSS protocol (39). **(C)** Cohort composition displayed as pie charts for laterality, choroidal invasion, optic nerve invasion, pT-category and *RB1* status.

The clinical and histopathological composition of the cohort is summarized in Figs. 1B,1C and Table S1. Cluster labels were assigned to the tumor samples based on Ryl et al. (*10*) (Cluster A (n = 3), Cluster B (n = 12) and Cluster C (n = 3). All Cluster A cases were female, while the sex distribution was balanced within Clusters B and C (Fig. 1B). There was variation in age at diagnosis across the clusters, with Cluster C patients being the youngest with a median age of 13 months at diagnosis (Interquartile Range (IQR) 3-33) and Cluster B patients being the oldest (31 months, IQR 21.25-45.75). Cluster A patients were diagnosed at a median age of 21 months (IQR 15-26). Two patients presented with bilateral retinoblastoma (P8 in Cluster B and P16 in Cluster C), while all other retinoblastomas occurred unilaterally (Fig. 1B and C). P1 underwent chemotherapy prior to enucleation, all other patients underwent primary enucleation (Fig. 1B). Minor choroidal invasion was observed in two Cluster B tumors (C1), while all remaining tumors were choroid-negative (C0) (Fig. 1C). No optic nerve invasion was observed in five tumors (N0), pre- or intralaminar invasion in 11 tumors (N1) and retrolaminar invasion in two tumors (N2) with clear surgical margins. Scleral infiltration was absent in all tumors (S0). Notably, one Cluster C tumor was *RB1^wt/wt^* (P17, Fig. 1B and C). Together, these comprehensive, matched genomic and epigenomic datasets provide a robust basis for the in-depth comparison of AH- and tumor-derived molecular profiles, as well as for methylation-based subtype assignment across the cohort.

### AH yields exceptionally high cfDNA amounts and ctDNA fractions

To establish AH liquid biopsies, we first assessed the cfDNA yields necessary for subsequent analysis, independent of patient or tumor characteristics and preanalytical limitations such as limited sample volumes. Collected AH sample volumes were very small, ranging from 2 µL to 251 µL (Fig. 2A, Cluster A 202.7 ± 6.8 µL, Cluster B 138.8 ± 79.18 µL, Cluster C 101.00 ± 85.28 µL). Nonetheless, high cfDNA abundance in AH allowed us to isolate sufficient amounts of cfDNA in all patients, with the exception of P1, who underwent secondary enucleation after chemotherapy (Fig. 2B, p=0.88). In this particular case, no cfDNA was detectable. Cell-free DNA purity, calculated as the proportion of cfDNA in total DNA exceeded 90% across all samples and clusters (Fig. S1A and Table S2, Cluster A 95.53 ± 1.85% Cluster B 100.60 ± 2.61%, Cluster C 97.81 ± 1.86%). Importantly, the estimated ctDNA fraction derived from WGS was high across the entire cohort with a median of 0.65 (IQR 0.44 – 0.93), supporting robust tumor detection (Fig. 2C and Table S2). ctDNA fraction did not differ significantly between clusters (p=0.85) and ctDNA was detected even in the sample from P1 cfDNA amounts below the limit of detection (Cluster A). This highlights the sensitivity of our liquid biopsy approach by amplifying dismal cfDNA input amounts, even below the limit of detection, to generate molecular ctDNA data. Consistently, ctDNA fraction was independent of AH input volume or cfDNA concentrations (Fig. S1B), and clinical and pathological parameters such as sex, age at diagnosis, or histopathological risk features (Fig. S1D).

**Figure 2.**
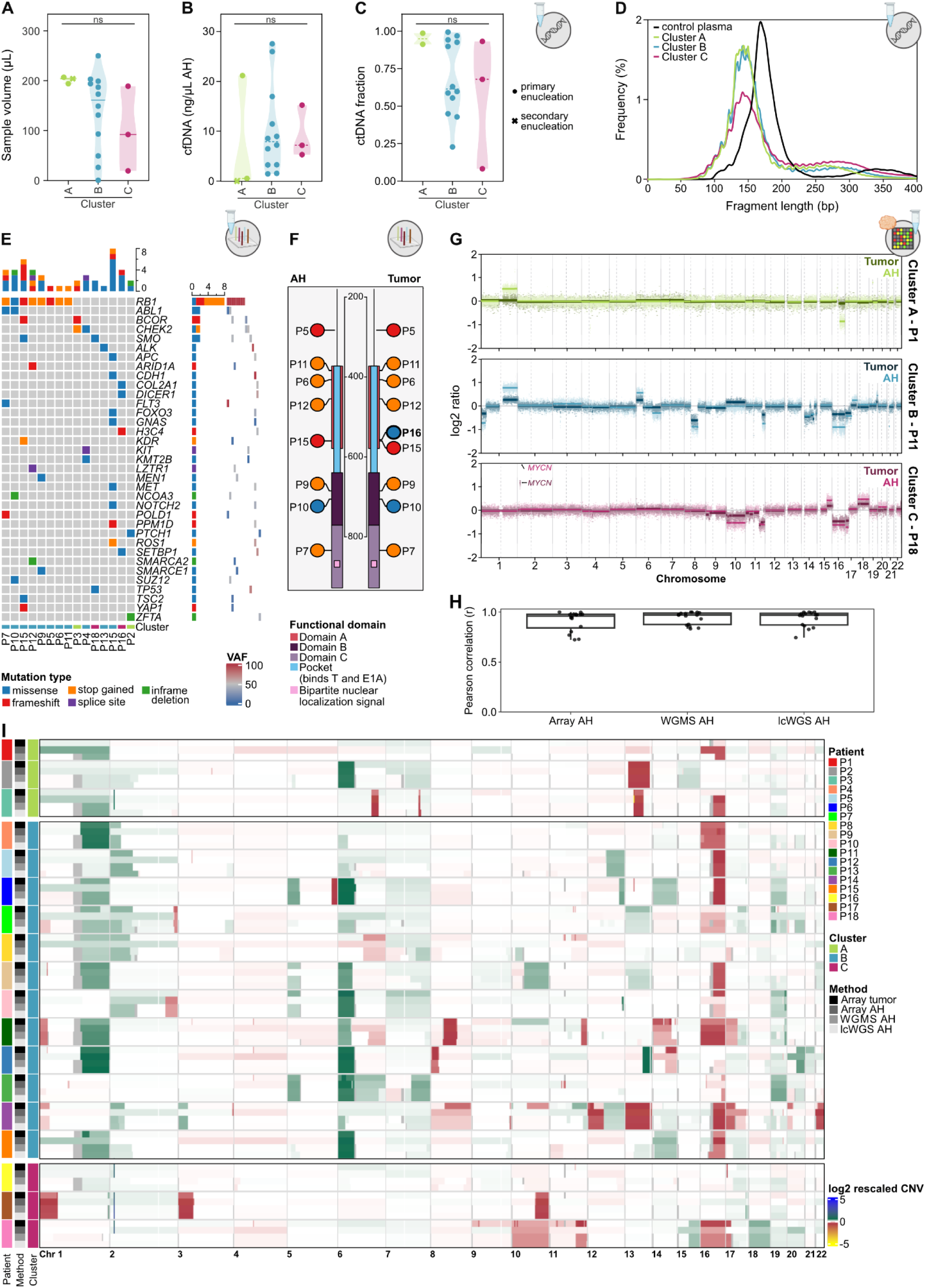
Reflection of Tumor biology in AH cfDNA through copy number, fragmentation, and mutation signatures. Violin plots showing **(A)** AH sample volume (p=0.27), **(B)** cfDNA concentration (p=0.88), and **(C)** ctDNA fraction (p=0.34) derived from lcWGS data, each stratified by retinoblastoma cluster. Violin color indicates cluster; point shape denotes enucleation type. Each dot represents one sample. **(D)** Fragment length density plots of AH-derived cfDNA from lcWGS, showing cluster-wise mean profiles for clusters A–C compared with control plasma. **(E)** Oncoprint summarizing SNVs detected in AH-derived cfDNA using targeted panel sequencing, with genes ordered by alteration frequency, mutation types color-coded, and variant allele frequencies shown as a heatmap. **(F)** Lollipop plot of *RB1* mutations detected in tumor DNA (right) and AH-derived cfDNA (left) by panel sequencing, colored by mutation type and mapped to RB1 functional domains. **(G)** Exemplary CNV plots of copy number alterations across retinoblastoma clusters A–C, derived from methylation arrays, overlayed for tumor (dark) and AH (light) samples. **(H)** Boxplots of patient-wise Pearson correlations between rescaled CNV profiles from AH cfDNA and matched tumor tissue, stratified by analytical method; correlations were computed using autosomal bins with informative CNVs. **(I)** Heatmap displaying rescaled log₂ CNV profiles across chromosomes 1–22, derived from methylation array (tumor and AH), WGMS (AH), and lcWGS (AH), annotated by patient, method, and cluster.

*In silico* assessment of paired-end sequenced DNA segments revealed a median cfDNA fragment length between 133 and 152 bp, which is substantially shorter than the ∼166 bp peak typically observed in healthy cfDNA plasma samples (Fig. 2D, Fig. S1C). Furthermore, we observed the previously described 10bp periodicity consistent with the protection observed for mono- and di-nucleosomes. The median fragment lengths were comparable across clusters and did not significantly differ by clinical covariates, including choroidal or optic nerve invasion and *MYCN* amplification status (Fig. S1E). Determining whether these patterns reflect general characteristics of AH or are specific to retinoblastoma will require additional validation against AH samples from non-oncological patients and those with other ocular malignancies. Collectively, these results indicate that liquid biopsies from AH reliably result in high-yield and - purity cfDNA with consistently detectable ctDNA fractions and characteristic nucleosomal fragmentation patterns, irrespective of clinical and histopathological characteristics. Most importantly, the presented work flow is not constrained by limited sample volumes.

### High confidence calling of single nucleotide variants and CNVs is achieved in AH ctDNA

For diagnostic purposes, it is critical to identify tumor-specific genetic alterations with predictive relevance. Therefore, we investigated which genomic and epigenomic alterations could be reliably recovered from AH-derived cfDNA. Targeted panel sequencing of AH cfDNA demonstrated the ability of liquid biopsies to capture retinoblastoma single nucleotide variants (SNVs). Alterations were identified in *RB1*, *ABL1*, *BCOR*, *CHEK2* and *SMO*, along with additional variants in genes implicated in chromatin remodeling and DNA damage response (Fig. 2E and Table S3). Across the cohort, the median variant allele frequency (VAF) of detected mutations was approximately 50%, with approximately 24% of the mutations exceeding 90%. Given that no matched constitutional DNA was available, we cannot distinguish somatic from germline variants; however, VAFs around 50% may reflect germline heterozygosity. In contrast, the very high VAFs observed for *RB1* alterations (96–100%) indicate loss of heterozygosity of the second allele. SNVs in *RB1* were detected in eight out of the nine cfDNA samples for which *RB1* SNVs were also detected in the tumor (Fig. 2F and Table S3).

Furthermore, AH Array derived genome-wide CNV profiles closely matched tumor profiles across all clusters as shown in representative patients (Fig. 2G). Canonical RB-associated genomic alterations, such as gains of 1q and 6p and loss of 16q, were reliably detected in both AH and matched tumor samples. Moreover, cluster-specific CNV signatures, as delineated by Ryl et al. (*10*), were faithfully reproduced: Cluster A was characterized by a 1q gain detectable only in AH, but not in the corresponding tumor (Fig. 2G); Cluster B by 1q and 6p gains together with 1p and 16q losses; and Cluster C by 1p loss, 2p gain, and 16q loss in both AH and tumor (Fig. 2G, Fig. S1F). For Cluster C patients, *MYCN* amplifications were detected in all three matched tumor and AH samples (Fig. 2G, Fig. S1F). Additional CNVs were observed in individual cases, reflecting inter-patient heterogeneity, yet overall demonstrating substantial concordance between AH- and tumor-derived profiles. Interestingly, the CNV profile of the *RB1^wt/wt^* samples showed a distinct pattern with an additional loss of 1p and 3p, which were not observed in the other Cluster C samples (Fig. S1G).

As AH cfDNA accurately reflects tumor CNV patterns, and because prior AH cfDNA studies in retinoblastoma have been restricted to a single analytical platform, this study was designed to comprehensively evaluate diverse cfDNA analysis methods and identify the most informative approach for tumor detection in AH cfDNA. For this purpose, all AH samples were analyzed by methylation array, lcWGS (except AH from P1), and WGMS, with matched tumor tissue profiled by methylation array as reference. To enable quantitative comparison across platforms, CNV amplitudes from all technologies were harmonized using a unified 1-Mb binning and rescaling framework, which enabled cross-platform harmonization of CNV signal amplitudes. Method-specific scale factors indicated higher CNV amplitudes for WGMS (1.97) and lcWGS (1.96) relative to Array-derived tumor profiles (reference = 1), while AH array-based CNVs showed a modest increase in amplitude (1.14) (Table S4). Correlation analysis of AH cfDNA and tumor DNA revealed high concordance between matched patient samples across all applied methods when restricting the analysis to informative genomic regions, defined by an absolute tumor log2 copy-number ratio greater than 0.2. Patient-wise correlations showed consistently high agreement for all platforms, with sequencing-based methods exhibiting slightly higher correlation coefficients (Fig. 2H; Array: median r = 0.96, mean r = 0.91 ± 0.10; WGMS: median r = 0.97, mean r = 0.94 ± 0.06; lcWGS: median r = 0.97, mean r = 0.93 ± 0.08), while patient-wise correlation coefficients could not be calculated for P1 and P16 due to insufficient numbers of informative tumor CNV bins resulting from largely flat tumor CNV profiles (Table S4). Pooled scatterplots across all informative patient–bin pairs further illustrated the high global concordance between AH-derived and tumor CNV profiles (Fig. S1H). Across all platforms CNVs were reliably detected and showed high intra-patient concordance (Fig. 2I). Even in AH from P1, where cfDNA was below the detection limit, CNV profiles could still be obtained, indicating that all applied approaches were capable of recovering CNV signals despite minimal input. MYCN amplification was detectable in the array-based data and, in selected cases, also in WGS-based profiles; however, for some patients MYCN gain was represented by a single 1-Mb bin and therefore did not form a contiguous CNV segment in the WGS-based segment files used for cross-platform harmonization. As a consequence, MYCN amplification was not consistently included in the segment-based comparative analyses, despite being visible at the bin level. Irrespective of the applied methods, some CNVs were only observed in the cfDNA but remained undetected in the tumor tissue, providing additional biological insight in at least three cases. This included a partial 2q loss in P10, detectable exclusively in AH cfDNA. Similarly, in P7 canonical alterations (6p gain and 16q loss) were absent in both tumor DNA and AH methylation array data but became detectable in lcWGS-AH and WGMS-AH profiles. These observations emphasize that higher-resolution sequencing-based methods enhance CNV detection sensitivity in samples with limited cfDNA input, thereby reducing the risk of false-negative results when using lower-resolution platforms. A more pronounced example is P3, which displayed a 1q gain and 16q loss in AH cfDNA that were not detectable in the time-matched tumor specimen. The clinical imaging supported a multifocal disease architecture, with multiple intravitreal and subretinal seeds visible on RetCam® examination (Fig. S1I). Given the presence of these tumor seeds and the absence of a germline *RB1* variant, these alterations likely reflect spatial heterogeneity arising from a pseudo multifocal tumor architecture rather than a hereditary predisposition. This illustrates how AH liquid biopsy can capture molecular features contributed by anatomically distinct tumor foci that may not be represented in a single tissue section. Notably, this patient also presented with *MYCN* gain in tumor and AH, despite not belonging to Cluster C.

Together, these results establish AH-derived cfDNA as a reliable surrogate for the genomic architecture of retinoblastoma, capturing both characteristic CNVs and high-confidence SNVs with strong concordance to tumor tissue. Collectively, all cfDNA analysis methods were able to reveal characteristic CNV patterns, with WGS-based approaches offering superior sensitivity and showing more potential for ultra-low input samples.

### Cluster-specific methylation patterns are preserved in AH cfDNA

With methylation classification becoming integral aspects of the latest WHO classifications for brain tumors and pediatric tumors (*36*), specific methylation patterns have also defined retinoblastoma subtypes associated with severity (*10*). Hence, we assessed whether these patterns are preserved in AH cfDNA enabling pretreatment patient stratification. Unsupervised embedding by t-distributed stochastic neighbor embedding (t-SNE), based on CpG sites previously identified by Ryl et al. (*10*) to distinguish retinoblastoma Clusters A, B, and C, demonstrated that AH samples grouped closest with their matched tumor tissues, and cluster assignments were preserved irrespective of biomaterial types (Fig. 3A). Only the samples from P3 strayed slightly with the AH clustering more closely with other Cluster B samples (Fig. 3A, teal circle and triangle). Upon incorporation of additional tumor and ocular control samples from independent cohorts (*9, 10, 37*), AH-derived profiles from our study demonstrated direct alignment with established retinoblastoma methylation groups. Overall, these profiles exhibited a clear distinction of retinoblastomas from normal retina and other ocular pathologies (Fig. S2A). Two normal neuroretina and one neuroretina in non-proliferative diabetic retinopathy samples aligned with retinoblastoma clusters. Density plots of β-values at discriminative CpG sites confirmed the anticipated hypermethylation signature in Cluster A and Cluster B, whereas Cluster C showed the expected hypomethylation pattern (Fig. 3B). These signatures exhibited analogous distributions between AH and tumor samples, (Fig. 3B). Subsequent analysis of individual AH samples further confirmed consistent cluster-specific patterns across patients (Fig. S2B). Heatmap visualization of CpG sites further supported the distinct separation of Cluster A and B hypermethylation patterns and Cluster C hypomethylation patterns across both AH and tumor samples (Fig. S2D). Given the discordant methylation signatures between tumor and AH of P3 in the t-SNE, this patient’s density plots were analyzed separately, revealing a Cluster A signature in the tumor and a Cluster B signature in the AH cfDNA (Fig. 3C). This divergence mirrors the CNV discrepancies observed in P3 (Fig. 2H) and is consistent with a suspected pseudo multifocal disease architecture (Fig. S1I), indicating that AH cfDNA captures molecular contributions from tumor foci not represented in the analyzed tissue section. Despite the MYCN amplification observed in the genomic analyses, there is distinct discrimination from Cluster C with regard to the methylation signature, as demonstrated by evident hypermethylation at Cluster C-specific CpGs.

**Figure 3.**
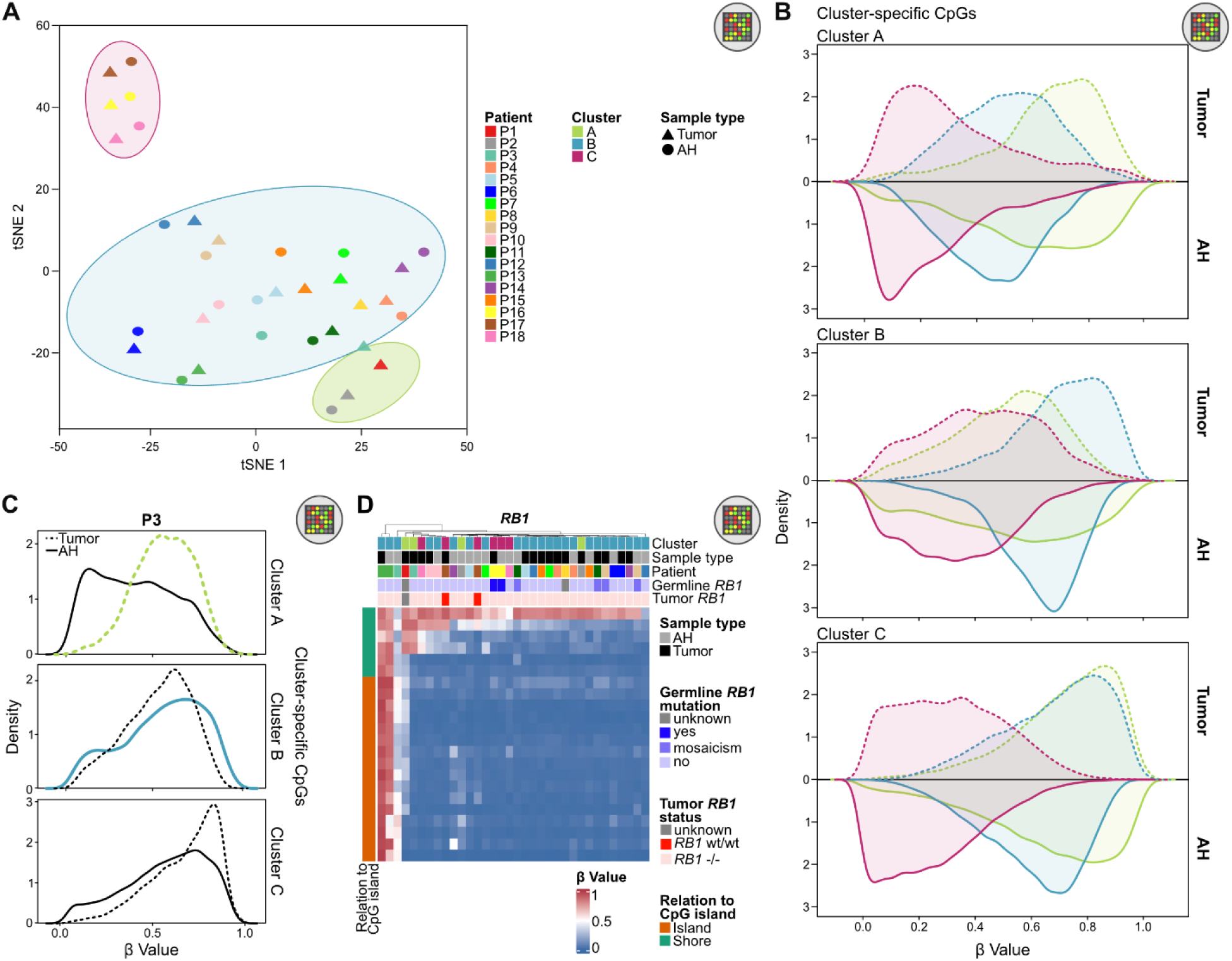
Cluster-specific methylation patterns in AH cfDNA. **(A)** t-SNE plot of tumor and AH samples based on CpG sites identified by Ryl et al. as discriminative between retinoblastoma clusters A, B, and C. Points are color-coded by patient and shaped by sample type (tumor vs. AH); ellipses indicate cluster assignment. **(B)** Density plots of β-values across cluster-discriminating CpG sites identified by Ryl et al., displayed by retinoblastoma cluster (color-coded) and sample type (tumor vs. AH). β-values are derived from methylation array profiles. **(C)** Density plots of β-values across cluster-discriminating CpG sites displayed for patient 3 (P3). Tumor (dotted) and AH (solid line) samples are overlayed. **(D)** Heatmap of β-values for CpG sites within the *RB1* promoter region, based on methylation array data. Rows represent CpG sites; columns represent tumor and AH samples. Samples are annotated by patient, retinoblastoma cluster, sample type, germline *RB1* status, and tumor *RB1* status.

The application of correlation analysis to AH-tumor pairs yielded the confirmation that samples were primarily clustered according to patient and cluster assignment, as opposed to tissue type (Fig. S2C). Pearson correlation between matched AH and tumor profiles, calculated on the selected CpGs reported by Ryl et al.(*10*), was uniformly high (mean r = 0.80, median r = 0.83). Notably, 9 of 15 patients (60%) showed correlations ≥ 0.8. Among these 15 patients, 11 AH–tumor pairs appeared as each other’s nearest neighbors in the correlation heatmap, whereas four pairs exhibited weaker intra-pair similarity. Taken together, this highlights the feasibility of reproducing cluster-defining methylation signatures in liquid biopsies. P1 and P8 were excluded due to low-quality array profiles, and P3 was excluded because its tumor and AH sample clustered inconsistently (Fig. 3C).

Furthermore, these findings demonstrate that cluster-specific methylation patterns originally described in tumor tissue are preserved in AH cfDNA, thereby enabling assignment of patients’ malignancies to molecular subtypes directly from liquid biopsy.

### In depth methylation analyses of AH cfDNA reveals *RB1* promoter inactivation and invasion-associated signatures

Next, we examined the promoter methylation signatures of selected genes that have been reported to play a role in retinoblastoma biology or tumor progression. Analysis of the *RB1* promoter region revealed consistent methylation profiles between the tumor and AH, with a single case (P13) showing clear promoter hypermethylation in both (Fig. 3D). Notably, this patient did not harbor genetic *RB1* mutations in tumor DNA or cfDNA, suggesting the promoter hypermethylation as *RB1* inactivation event. Biallelic promotor hypermethylation has been identified as pathogenic *RB1* alteration in tumor diagnostics (*18*). To our knowledge, this is the first demonstration of the feasibility to detect *RB1* promoter hypermethylation in cfDNA.

Furthermore, we performed in-depth analyses of the promoters of *NHLH1*, *NTN3* and *TFF1,*, genes linked to invasive retinoblastoma phenotypes (*9, 10*). *NHLH1*, a helix–loop–helix transcription factor, and *NTN3*, a netrin family member involved in axon guidance and cell migration, have been associated with proliferative and invasive phenotypes (*10*). In contrast, *TFF1* (trefoil factor 1) is associated with epithelial cancers and plays a role in invasion and cellular survival signaling (*9*). Ridge plots revealed distinct methylation distributions with the optic nerve invasion categories N0, N1 and N2 (Fig. 4A): N0 denotes absence of invasion; N1, pre- or intralaminar invasion; and N2, retrolaminar invasion with tumor-free resection margin. In both tumor and AH samples, we observed more pronounced promoter hypomethylation with increasing optic nerve invasion, which is consistent with an association between reduced methylation and more aggressive disease. Heatmaps of individual CpG sites confirmed these patterns at single-CpG site resolution, illustrating reproducible promoter hypomethylation changes in both tumor and liquid biopsy (Fig. S3A).

**Figure 4.**
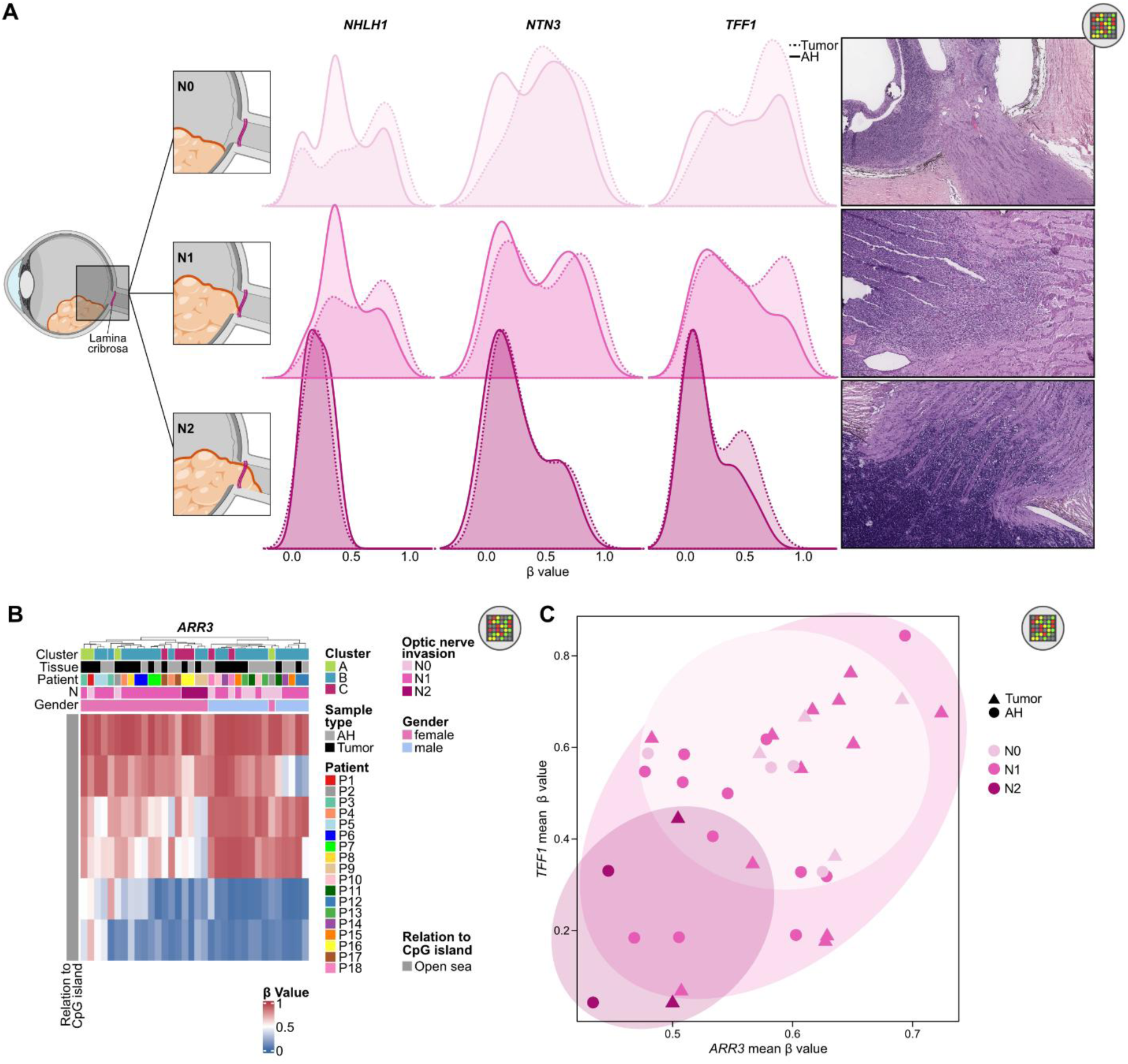
Integrative CNV and promoter methylation analysis across methods and clinical subgroups in retinoblastoma. **(A)** Ridge plots of β-values across promoter regions of *NHLH1*, *NTN3*, and *TFF1*, based on methylation array data. Tumor (dotted) and AH (solid line) samples are overlayed, and ridges are stratified by optic nerve invasion status (N0–N2) from top (N0) to bottom (N2) **(B)** Heatmap of β-values for CpG sites within the *ARR3* promoter region, based on methylation array data. Rows represent CpG sites; columns represent tumor and AH samples. Samples are annotated by patient, retinoblastoma cluster, sample type, optic nerve invasion and gender. **(C)** Scatterplot of mean promoter β-values for *ARR3* (x-axis) and *TFF1* (y-axis). Points are colored by optic nerve invasion status (N0–N2) and shaped by sample type (tumor vs. AH).

A poor prognosis has been shown to be associated with the expression of *ARR3* in combination with *TFF1* (*38*). Heatmap analysis of ARR3 promotor methylation showed an association between hypomethylation and optic nerve invasion (Fig. 4B). However, since *ARR3* is located on the X chromosome samples clustered mostly based on gender. Correlation of mean ß values of *TFF1* and *ARR3* promoters showed more invasive tumors correlating with non-mutually exclusive hypomethylation of both gene promotors (Fig. 4C).

Together, these results demonstrate that promoter methylation signatures in AH cfDNA closely mirror those in tumor tissue, capturing clinically relevant epigenetic changes, including *RB1* inactivation through hypermethylation and invasion-associated hypomethylation events in *NHLH1*, *NTN3*, *TFF1*, and *ARR3*. Notably, *NHLH1*, *NTN3*, and *TFF1* represent commonly reported protein biomarkers of retinoblastoma invasion, highlighting the potential of cfDNA methylation profiling to enable multi-biomarker extraction from a single assay and thereby support minimally invasive patient-stratification strategies.

### A novel XGBoost machine-learning classifier is established for retinoblastoma liquid biopsies

In order to enhance clinical applicability of our findings, we aimed to develop a classifier able to determine retinoblastoma clusters from methylation signatures from minimally invasive AH samples (Fig. 5A). For balanced cohort composition, we incorporated datasets published by Ryl et al. (RB) (*10*) and Berdasco et al. (normal retina, ocular disease control) (*37*) to increase sample size and robustness of the classifier. In total, the combined training dataset consisted of 114 samples, including 35 Cluster A, 29 Cluster B, 13 Cluster C, 17 disease controls, and 20 normal retina samples. The disease control class included fibrovascular membranes from proliferative diabetic retinopathy and membranes from proliferative vitreoretinopathy (Fig. S2A). Comparison of AH- and tumor samples revealed differentially methylated CpGs, which were subsequently excluded from the classifier as they could make the classifier’s predictions dependent on the sample type. Consequently, the classifier is sample-type agnostic (Fig. 5B and Table S5). To reduce dimensionality, the DNA methylome was restricted to the 100,000 most variable CpG sites across all samples. Separately, a one-versus-rest limma analysis was performed for each class to determine discriminative features, from which the top 2,000 differentially methylated CpGs per class were selected. (Fig. S4A and Table S6). Notably, a substantial proportion of Cluster C-specific CpGs were hypomethylated, aligning with the preceding analyses demonstrating that Cluster C is characterized by hypomethylation (Fig. 3A). After intersecting these class-discriminative CpGs with the variance-filtered matrix and excluding CpGs identified in the paired AH–tumor analysis, a final feature set of 8,999 CpGs was obtained (Table S7). A classifier was trained using gradient-boosted decision trees, with five-fold cross-validation for performance assessment (Table S8). The dataset from Liu et al. (*9*) was used for independent validation (Table S9).

**Figure 5.**
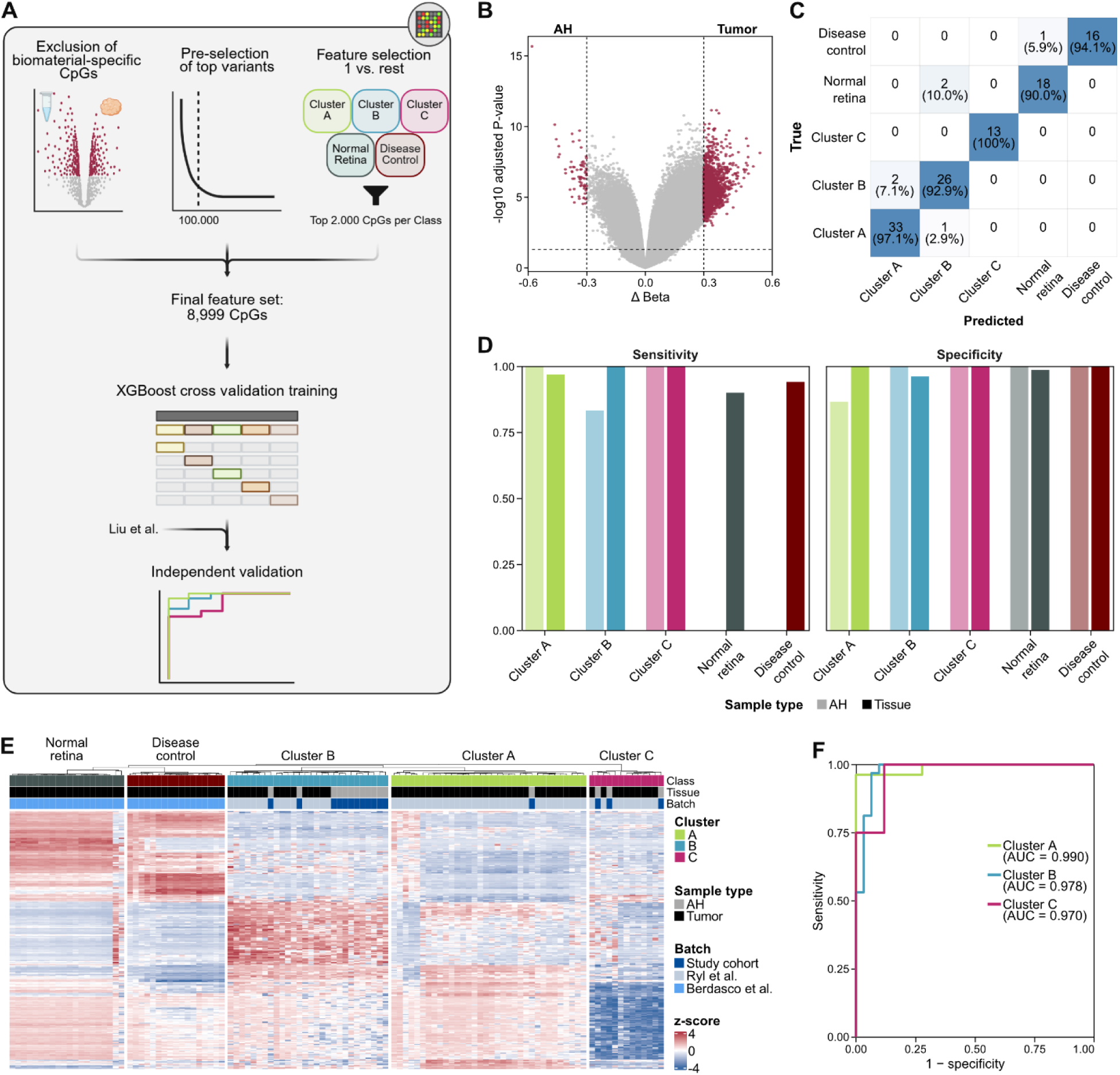
Machine learning-based classification of retinoblastoma subtypes using tumor and liquid biopsy methylation profiles. **(A)** Graphical overview of the classifier architecture. **(B)** Volcano plot of differential methylation analysis between matched tumor and AH samples, based on methylation array data. Analysis was performed using the limma framework on paired samples with thresholds of adjusted p < 0.05 and |log2 fold change| > 0.1. **(C)** Confusion matrix of the XGBoost classifier trained on methylation array data from both tumor and AH samples to distinguish between retinoblastoma clusters, normal retina, and disease control samples. Values indicate the number and percentage of samples per class predicted correctly or misclassified. **(D)** Comparison of sensitivity and specificity of the classifier detecting the individual classes in tissue DNA and AH cfDNA. **(E)** Heatmap of z-scores of the 300 most important CpGs. Rows represent CpG sites; columns represent tissue and AH samples. Samples are annotated by class, sample source and batch. **(F)** ROC curves for the classifier applied to the Liu et al. cohort for independent validation. Normal retina and disease control are not represented in the external dataset.

Following feature selection and cross-validation, overall accuracy of class prediction exceeded 94% across all classes (Fig. 5C). One disease control (5.9% of class) and two normal retina samples (10%) were misclassified as normal retina and Cluster B, respectively. The prediction of all Cluster C samples was correct. Two Cluster B samples (7.1%) were wrongly classified as Cluster A, whereas one Cluster A sample (2.9%) was classified as Cluster B. Importantly, no retinoblastoma samples were classified as normal retina or disease control (Table S8). Throughout the classes discriminatory power was high with area under the receiver operating characteristic curve values (AUC) ranging from 0.96 to 1.00 (Fig. S4E, Cluster A: 0.999, Cluster B: 0.985, Cluster C: 1, normal retina: 0.964, disease control: 0.998). Additionally, sensitivity and specificity were high and largely consistent across biomaterial source (Fig. 5D). Sensitivity for AH samples was over 80% for all Clusters, while specificity also exceeded 80% for all classes. It is noteworthy that for Cluster B and normal retina specificity regarding AH samples surpassed that of tissue.

Analysis of the 300 most important features of the classifier revealed CpGs that clearly differ between retinoblastoma and other classes as well as features showing distinct z-scores between the clusters (Fig. 5E and Table S10 for the full ranked list)). Five Cluster A samples and two normal retina samples showed a z-score signature that differed from all other samples of their respective class, which showed homogenous signatures. Informed t-SNE clustering based on the 300 most important features showed that the five Cluster A samples cluster closer to normal retina and disease control and the two normal retina samples cluster with Cluster A/B (Fig. S4B). The latter are the same that were wrongly predicted as Cluster B (Fig. 5C). The 30 overall most important CpGs as well as the two most important CpGs per class are shown in Fig. S4C and D.

Applying the classifier to the Liu et al. (*9*) cohort, we first assigned samples to retinoblastoma clusters based on the correspondence described by Ryl et al.(*10*), mapping subtype 1 to Cluster A, subtype 2 without *MYCN* amplification to Cluster B, and subtype 2 with *MYCN* amplification to Cluster C. This resulted in 63 validation samples, consisting of 27 Cluster A, 32 Cluster B, and 4 Cluster C cases. AUCs still exceeded 0.97 for all clusters showing the high discriminatory power and robustness of the classifier in an independent and unseen dataset (Fig. 5F and Table S9, Cluster A: 0.990, Cluster B: 0.978, Cluster C: 0.970). In addition, visualization of the top 300 most important CpGs from the final model in a correlation heatmap (Fig. S4G) showed that samples again clustered clearly according to their respective groups.

To evaluate the reliability of predictions, the highest predicted class score was analyzed for the cross-validation as well as the independent validation (Fig. S4F and Supplementary Tables 8 and 9). The majority of the correct predictions were made with high probability. Only one and two samples, respectively, were classified wrongly with high probability (over 0.9) and in each data set two samples were predicted correctly with a predicted probability of less than 0.5. Collectively, this indicates that predictions assigned a high probability are reliable.

Notably, when looking at the difference in sensitivity between cross- and independent validation, the sensitivity in the independent dataset for Cluster B exceeded the training data set, with all samples detected. The greatest difference can be seen for Cluster C were the independent validation only reached 0.5 with two samples misclassified as Cluster B (Fig. S4H). As Liu et al. did not differentiate into Cluster B and C, true cluster assignment was based on all *MYCN* being classified as Cluster C. However, density plots revealed that RB15 actually exhibited a methylation signature characteristic of Cluster B (Fig. S4I). This finding aligns with the classifier’s high probability prediction of Cluster B, which had a probability of 0.84. In the case of RB224, the highest probability class was found to be Cluster B, with a probability of 0.39, thus showing no definitive cluster assignment. Concordantly, manual ß value analysis of RB224 did not show any cluster-specific methylation patterns. Classifier assignment of RB14 and RB224 matched the true cluster (Cluster C). Specificity did not vary greatly between training and test datasets.

Calibration analysis assessed the agreement between predicted class probabilities and observed outcomes on the independent validation cohort. Overall probabilistic performance was acceptable, with a multiclass Brier score of 0.1205, indicating accurate probability estimates across classes. Class-wise one-versus-rest Brier scores further demonstrated differences in calibration precision, with the lowest Brier score observed for Cluster C (0.0237), followed by Cluster A (0.0439) and Cluster B (0.0498), indicating the most precise probability estimates for Cluster C. Reliability diagrams illustrating class-wise calibration behavior are shown in Fig. S4J. While overall calibration was acceptable, further improvements may be achieved with larger validation cohorts.

## DISCUSSION

This study provides the first comprehensive integration of genetic and epigenetic profiling from AH–derived cfDNA in retinoblastoma, establishing liquid biopsy as a promising avenue for both molecular classification and risk stratification. Unlike previous work limited to genetic analyses (*26–28*), our integrative approach expands AH cfDNA analysis to allow for methylation-based risk group stratification. The novel retinoblastoma methylation classifier demonstrates an unprecedented high sensitivity and specificity in the classification of risk clusters independent of available biomaterial.

This study has several limitations. Due to the low incidence of retinoblastoma, the cohort size was modest and limited to eyes post-enucleation. Validation of our findings in larger, prospective cohorts, that also include conservatively treated eyes, is needed. Furthermore, longitudinal sampling could establish whether AH cfDNA can monitor disease progression, treatment response or the development of therapeutic resistance. Initial studies suggest that AH cfDNA monitoring does correlate with treatment response (*27*).

Furthermore, one must consider, that AH liquid biopsy implementation may be hindered by the invasiveness of the procedure, the potential risk of tumor cell spread in advanced disease and the necessity of general anesthesia for AH sample collection, given the patients’ young age. Plasma liquid biopsies would be more accessible. However, studies have demonstrated the far superior detection of tumor-derived signatures in AH cfDNA compared to plasma cfDNA in retinoblastoma (*28, 39*). Additionally, patients with retinoblastoma frequently undergo general anesthesia for diagnostic or treatment reasons facilitating AH collection. Chigane et al. have previously demonstrated that anterior chamber paracentesis carries a low risk profile, with only a single minor complication observed among over 1,200 procedures (*35*). Importantly, no evidence of tumor cell spread was detected. Furthermore, AH liquid biopsies enable the precise allocation of findings to a specific eye in patients with bilateral retinoblastoma. Consequently, AH liquid biopsies are the best possible choice in retinoblastoma.

A key limitation in the establishment of retinoblastoma liquid biopsies is the limited availability of informative biomarkers in the circulation or the constrained AH volume available. Nevertheless, this study demonstrates the ability to isolate high yields of cfDNA and ctDNA from AH and allowed the orthogonal comparison of methodological platforms. This enabled a comprehensive comparison of epigenetic and genetic cfDNA assays to establish robust SNV, CNV and methylation detection for prospective clinical implementation and lays the fundamental basis for well-informed platform prioritization for limited material, such as scenarios prior to enucleation or pre-treated retinoblastoma tumors. Our multi-omics in-depth data illuminates multiple aspects of retinoblastoma biology, invasion, and evolution on a gene-specific level. Furthermore, all assays allow tumor agnostic analysis, facilitating their implementation in pre-enucleation care, where molecular tumor data is currently unavailable. The robust ctDNA fraction, with the median exceeding 65% across the cohort, regardless of clinical parameters or enucleation type, allowed us to detect not only canonical CNVs, such as gain of 1q and 6p or loss of 16q, but also to resolve cluster-specific CNV signatures with strong concordance between AH and tumor tissue. Similarly, SNV analysis of AH cfDNA captured recurrent driver alterations, most notably *RB1* mutations. Interestingly, in several cases AH liquid biopsy was able to detect additional alterations, possibly allowing representation of multiple tumor foci that may not be evident in a solitary tissue analysis. Furthermore, the reliable detectability of poor prognostic markers like *MYCN* amplification in AH cfDNA, highlights the potential for pre-enucleation detection of aggressive, therapy-resistant tumors. In turn, patients could be spared adverse effects and long-term toxicity associated with conservative retinoblastoma therapy.

Our study confirms findings from Li et al. (*28*) showing the possibility of reliable methylation signature analysis in AH cfDNA. In addition, by developing and applying the first methylation-based multi-class classifier, we extend these findings and take a decisive step toward clinical implementation. By carefully excluding AH-tumor differentials prior to training, we minimized tissue-specific bias and ensured that the model learned true disease-related signals by including negative control samples. Despite the modest size of our primary cohort, the integration of public retinoblastoma methylation datasets substantially enhanced the robustness and generalizability of our findings. From a clinical perspective, the ability to classify retinoblastoma tumors into methylation clusters without the necessity for tissue biopsy represents a significant advancement in the field of precision diagnostics. The classifier could inform treatment intensity, guide enucleation decisions, and ultimately reduce the risk of over- or undertreatment. Importantly, independent of biological specimen, the classifier reliably distinguished canonical retinoblastoma subgroups as well as non-retinoblastoma controls, suggesting potential utility in challenging diagnostic settings. The misclassification of two normal retina samples, both derived from adult tissue, likely reflects the limited availability of pediatric retinal reference profiles and illustrates that classifier performance for non-tumor classes may depend on age-appropriate control datasets. The reduced sensitivity for Cluster C observed in the independent validation cohort is on the one hand attributable to the very small number of available *MYCN*-amplified cases, underscoring the need for larger subtype-specific cohorts for future benchmarking. On the other hand, manual β value analysis revealed that the misclassified cases exhibited either an absence of clear cluster alignment with any cluster or a methylation pattern consistent with Cluster B, despite being *MYCN* amplified. This is in line with our findings in P3 that showed that not all *MYCN*-amplified retinoblastomas belong to Cluster C.

Overall, limitations include small cohort size, the unavailability of non-RB-AH controls as well as the absence of pediatric non-retinoblastoma tissue control cases inherently constrained in the pediatric setting. Therefore, AH and biopsy material should be systematically collected from children undergoing paracentesis, biopsy, or enucleation for other ocular pathologies. Future efforts will focus on classifier validation in prospective, pre-enucleation liquid biopsy cohorts. Ultimately, methylation-based classification will be complemented by CNV- and SNV-based cfDNA information and aid to position liquid biopsy as a cornerstone of precision diagnostics for retinoblastoma.

Our study provides only the second published case of AH-based evidence of *RB1* promoter hypermethylation - as an alternative mechanism of gene inactivation (*28*). This extends the molecular insights into tumor biology obtainable through liquid biopsy beyond CNVs and SNVs. Promoter methylation status of *NHLH1, NTN3*, *TFF1* and *ARR3* revealed epigenetic regulations are strongly associated with optic nerve invasion. These genes, which play a role in neuronal development, signaling and tumor progression, exhibited progressive hypomethylation as the invasion stage increased from N0 to N2. Notably, these changes were consistently detected in both tumor and AH cfDNA, demonstrating that AH liquid biopsy can capture progression-related epigenetic signatures associated with histopathological risk features. Taken together, our liquid biopsy data capture hallmarks of conventional retinoblastoma diagnosis including *TFF1* expression currently performed as ELISA protein assays, *MYCN* amplification currently performed by FISH (*40*) as well as extent of invasion both performed on tumor tissue sections from a single assay providing clinically relevant prognostic information (*41*).

In light of our study, AH cfDNA profiling has the potential to significantly improve retinoblastoma care by enabling earlier, safer, and more precise tumor characterization while facilitating the discovery of novel biomarkers. With further validation, integration of AH liquid biopsy into the diagnostic pathway could allow molecular classification and risk stratification prior to enucleation, guiding personalized treatment decisions and surveillance strategies. Longitudinal AH cfDNA monitoring may ultimately provide a minimally invasive means to track disease dynamics and treatment response, complementing clinical staging with actionable biological insight.

## MATERIALS AND METHODS

### Patients and sample collection

The cohort consists of 18 intraocular primary retinoblastoma samples and matched AH liquid biopsies from patients diagnosed and treated between April 1^st^ 2023 and June 10^th^ 2024. Selection criteria for this study were (i) written informed consent for research project available and (ii) availability of matched AH liquid biopsy and retinoblastoma tumor tissue. The study was conducted in accordance with the Declaration of Helsinki. The study was approved by the Ethics Committee of the University Duisburg-Essen (ethics approval ID: 22-10845-BO approved November 22^nd^ 2022 and 23-11503-BO approved January 08^th^ 2024).

For retinoblastoma liquid biopsies, AH samples were obtained from retinoblastoma eyes collected immediately after enucleation. The procedure was meticulously executed to circumvent the risk of contamination and with the utmost care taken to ensure that the samples were obtained in a sterile manner. Subsequent to collection, AH samples and tumor tissue were frozen and stored at -80°C until further processing.

*RB1* variants were identified using DNA from fresh-frozen tumor samples or DNA from blood and one or more of the following in routine diagnostics (Dept. of Human Genetics, Essen) as previously described (*18, 42–44*): analysis of allele loss in tumors, cytogenetic analysis, denaturing high performance liquid chromatography, exon-by-exon sequencing, multiplex ligation-dependent probe amplification, methylation-sensitive PCR, quantitative fluorescent multiplexed PCR, quantitative real-time PCR, real-time PCR and single-strand conformation polymorphism analysis. Histopathological staging of tumors was performed according to the International Retinoblastoma Staging System (IRSS) (*45*).

Clinical data were obtained from the binational RB-Registry, a prospective clinical registry that systematically collects epidemiological information and longitudinal outcomes of patients with retinoblastoma.

### CfDNA isolation from AH samples

Cell-free DNA from AH liquid biopsies was isolated using the QIAamp Circulating Nucleic Acid Kit (Qiagen, Hilden, Germany) in accordance with the manufacturer’s protocol. The isolated cfDNA was eluted in 25-30 µL elution buffer provided with the kit. Cell-free DNA concentration and potential genomic DNA contamination were assessed using the 2100 Bioanalyzer (Agilent, Santa Clara, CA, USA) with the High Sensitivity DNA Kit. Cell-free DNA quantification analyzed mono-, di- and tri-nucleosomal peaks in the range between 150–500 bp, while genomic DNA contamination was evaluated across 50–7,000 bp fragments. Cell-free DNA purity was calculated as proportion of cfDNA in total DNA.

### Low-coverage whole genome sequencing (lcWGS) of cfDNA

For the downstream analysis of cfDNA isolated from AH liquid biopsies, lcWGS was performed. Library preparation for lcWGS was conducted employing the Accel-NGS 2S Hyb DNA Library Kit (Swift Biosciences, Ann Arbor, MI, USA). The input cfDNA ranged between 500 and 4000 pg per sample, as determined by Bioanalyzer measurements. Libraries were amplified with 12 to 14 PCR cycles, with the incorporation of no-template controls (unique molecular identifiers (UMIs); Swift Biosciences, Ann Arbor, MI, USA) to ensure the accuracy of the results. The quantification of libraries was conducted using the Bioanalyzer (Agilent, Santa Clara, CA, USA), after which they were pooled in equimolar ratios for sequencing on NovaSeq6000 or NovaSeqX 25 b 150 PE (Illumina, San Diego, CA, USA) with a target coverage of 3×.

### DNA methylation profiling of cfDNA and tumor DNA

To analyze DNA methylation patterns in AH liquid biopsies, methylation profiling was enabled by applying the NEBNext Enzymatic Methyl-seq Kit (New England Biolabs, Ipswich, MA, USA), with cfDNA target input amounts of 500-2000 pg. Polymerase chain reaction (PCR) amplification was conducted to enrich the libraries with 12-14 cycles. Following library preparation, the methylation libraries were quantified using Bioanalyzer (Agilent, Santa Clara, CA, USA). Array-based DNA methylation profiling was performed on the Infinium MethylationEPIC v2.0 BeadChip arrays (Illumina, San Diego, CA, USA), in accordance with the manufacturers protocol. Sequencing-based methylation profiling of cfDNA, was executed after equimolar multiplexing and subsequent sequencing on NovaSeqX, 25b 150 PE (Illumina, San Diego, CA, USA) with a target coverage of 3×. DNA methylation data from tumor biopsies were generated via the Infinium MethylationEPIC v2.0 BeadChip array (Illumina, San Diego, CA, USA), used according to the manufacturer’s protocol. DNA input ranged between 200 and 2000 ng.

### Panel Sequencing of cfDNA

For robust detection of SNVs and small insertions and deletions (indels) indels, targeted panel sequencing was performed on cfDNA from AH liquid biopsies. Libraries were prepared using the SureSelect XT HS2 DNA Reagent Kit (Agilent). The input cfDNA ranged between 500 and 20000 pg per samples, as determined by TapeStation measurements using the HS D1000 assay (Agilent, Santa Clara, CA, USA). Libraries were amplified prior to hybridization capture with 12 to 15 PCR cycles incorporating dual molecular barcodes (MBCs) for accurate deduplication in downstream analysis. Subsequently, hybridization capture of target regions was carried out using SureSelect Custom DNA Target Enrichment Probes (Agilent, Santa Clara, CA, USA). The target regions of the custom panel comprised all coding regions of 191 genes relevant to CNS tumors, along with selected intronic and promoter regions, for a total capture region size of ∼1.3 Mb. Post-capture libraries were again amplified with 11 to 12 PCR cycles and pooled in equimolar ratios for paired-end sequencing on a NovaSeq X Plus instrument using 1.5B (200 cycles) Kits (Illumina, San Diego, CA, USA). The aimed raw on-target coverage was 15,000× for AH cfDNA samples.

### Bioinformatics Analysis

#### Low-coverage whole-genome sequencing (lcWGS)

Initial processing of lcWGS data was conducted utilizing the in-house Omics IT and Data Management Core Facility’s (ODCF) proprietary AlignmentAndQCWorkflows (v1.2.73-1) (*46*). The workflow encompassed adapter trimming, alignment of sequences to the human reference genome GRCh37/hg19 (*47*), marking of duplicate reads, sorting, and extraction of sequence quality metrics. The unique molecular index (UMI) processing was conducted using the fgbio workflow (v1.1.0, Fulcrum Genomics) and the Picard toolkit, enabling BAM file collapse and the generation of high-quality, de-duplicated molecular consensus reads. Subsequent to this, the reads were realigned using BWA-MEM (*48*).

In the context of copy number and fragmentomic analysis, CNV segmentation was performed using ichorCNA (v0.3.2) (*49*), based on normalized log2 ratios within 1 Mb genomic windows. As cfDNA of AH may contain high fractions of ctDNA, ichorCNA default settings, originally optimized for plasma samples with lower tumor fractions, were adjusted to better accommodate the tumor content observed in AH samples as follows:–ploidy “c(2)”–normal “c(seq(0.2,0.8,0.1),0.9)”–maxCN 5–includeHOMD FALSE–chrTrain “c(1:18,20:22)”–estimateNormal True–estimatePloidy True–estimateScPrevalence True–minSegmentBin 10–altFracThreshold 0.05–scStates “c(1,3)”. Fragment length profiles were derived using the cfdnakit package (*50*), with cfDNA fragment lengths determined from paired-end sequencing reads based on the genomic coordinates of both read ends after alignment.

#### Panel Sequencing

The targeted panel sequencing data was analyzed using an in-house pipeline based on the Illumina DRAGEN Secondary Analysis platform (Illumina, San Diego, CA, USA). The raw sequencing output was further processed (demultiplexing, trimming, deduplication, alignment/mapping to the human reference genome GRCh38, variant calling) by the DRAGEN software v4.4 and the Picard toolkit was used for quality control. Called variants were annotated using VEP and SnpEff. Downstream filtering was applied to only include calls that occur in at least two de-duplicated consensus reads at positions with at least 8x coverage and to remove variants with allele frequencies below 3% and a gnomAD frequencies above 0.1 %. On a biological level, variants were filtered to further exclude synonymous variants without predicted splice effect, deep intronic variants (±15 bp exon-intron boundary) and noncoding variants in 3’/5’_UTRs of the target genes (except for the *TERT*-promoter region).

#### DNA Methylation Profiling (Array)

DNA methylation profiling was performed using the Illumina Infinium MethylationEPIC v2.0 BeadChip platform. Raw IDAT files were processed in R (v4.1.3) with the minfi package (*51*), applying background correction and dye-bias normalization using the preprocessNoob function. Copy number variations (CNVs) were inferred from array intensity data using the conumee package (*52*). Heatmaps of β-values and CNV profiles were generated with ComplexHeatmap (*53, 54*). For large-scale t-SNE embeddings including additional samples from Liu et al. (*9*), Ryl et al. (*10*), and Berdasco et al. (*37*), batch effects were corrected using the sva package (*55*) with the ComBat() function prior to dimensionality reduction. t-SNE analysis was performed with the Rtsne package (*56*).

Methylation density distributions were calculated with minfi (*51*), and missing values were imputed using the impute package with the impute.knn() function. Ridge plots of promoter methylation patterns were generated with ggridges (*57*), and all other visualizations were created with ggplot2 (*58*).

#### DNA Methylation Profiling (WGS)

EM-seq data were processed with the nf-core/methylseq pipeline (v2.6.0) (*59*) using Bismark for alignment to the GRCh38 reference genome (*60*). Reads were adapter-trimmed with TrimGalore! (*61*), quality assessed with FastQC (*62*) and MultiQC (*63*), and duplicate reads were removed with the Bismark deduplication module. Libraries were processed in EM-seq mode (--em_seq true), and CpG methylation calls were generated with the Bismark methylation extractor (--comprehensive). CNVs and tumor fractions were inferred from EM-seq read depth using the cfdna Python library (*64*). For the cross-platform CNV comparison (Array, lcWGS, EM-seq), EM-seq data were additionally re-aligned, using the same ODCF in-house alignment workflow (*46*) as lcWGS, to the GRCh37/hg19 (*47*) reference. These hg19 alignments were used exclusively for CNV segmentation to ensure a uniform genomic reference across platforms and to avoid inaccuracies associated with liftover-based CNV comparison. All methylation analyses continued to use the primary Bismark/GRCh38 workflow.

#### Cross-platform CNV harmonization and correlation

To enable quantitative comparison of CNV profiles across Array, lcWGS, and EM-seq, segment-level CNV calls from all platforms were mapped to the QDNAseq 1 Mb genome annotation (hg19) (*65*). For each sample, overlapping segments were aggregated within each bin by averaging their segment medians. All samples were then independently centered by subtracting their autosomal (chr1–22) median log2 value. Genomic interval operations and overlap computations were performed using the GenomicRanges (*66*) and rtracklayer (*67*) packages. For each patient with matched tumor and AH samples, autosomal bins were paired between Array Tumor and each AH method. Only bins with clearly interpretable CNVs were used (|Array Tumor log2| ≥ 0.2 and |AH log2| ≥ 0.05, same direction of change). For each patient–method pair, a scale factor was derived as the median ratio between AH and Array Tumor log2-centered values. Method-specific global scale factors were derived as the median of the patient-level slopes, providing a single rescaling constant per technology. Array Tumor served as the reference with a fixed scale factor of one. All CNV profiles were rescaled by dividing the centered log2 values by the corresponding platform-specific factor, producing harmonized CNV amplitudes across technologies.

To assess concordance between tumor tissue and AH-derived CNV profiles, Pearson correlation coefficients were calculated using the rescaled log2 values. Correlations were computed on autosomal bins only and restricted to informative genomic regions, defined as bins with an absolute log2 copy-number ratio greater than 0.2 in the tumor sample. Correlations were calculated on a per-patient basis for each AH method, and summarized across patients using boxplots. In addition, pooled correlations across all informative patient–bin pairs were visualized using scatterplots to illustrate global cross-platform agreement.

#### Methylation-based XGBoost classifier

We developed a methylation-based multi-class classifier to assign samples to retinoblastoma clusters A, B, and C as well as “Normal Retina” and “Disease Control” based on Illumina array β-values. Since our primary cohort consisted of only 18 patients, we extended the dataset with published methylation data from Ryl et al. (*10*) and Berdasco et al. (*37*), ensuring sufficient representation of all classes by assembling a total of 114 samples, including 16 AH and 98 tissue specimens. This combined dataset comprised 35 Cluster A samples (34 tumor, 1 AH), 29 Cluster B samples (12 AH, 17 tissue), 13 Cluster C samples (3 AH, 10 tissue), 17 disease control samples consisting of fibrovascular membranes from proliferative diabetic retinopathy and membranes from proliferative vitreoretinopathy, and 20 normal retina samples. Arrays were normalized per batch using preprocessNoob from the minfi package (*51*) and merged into a common matrix restricted to CpG probes present across all batches. To reduce dimensionality, we retained the 100,000 most variable CpGs (row variance of β-values, matrixStats).

To avoid the classifier learning systematic differences between aqueous humor (AH) and tumor tissue, we first performed a paired differential methylation analysis on patients with matched AH–tumor samples using the limma package (*68*) (design: ∼ patient + tissue, AH as reference; empirical Bayes moderation with eBayes (robust=TRUE)). CpGs with adjusted P < 0.01 and |logFC| > 0.3 were identified. Importantly, the 18 matched tumor samples were used only for this filtering step and not included in model training or cross-validation.

Feature selection for classification was then carried out in a one-versus-rest design using limma with equal-class contrasts. For each target class, we compared the class against the average of all others and ranked CpGs by adjusted P value and logFC. The top 2,000 CpGs per class were selected, and the union across classes yielded a fixed feature set. After intersecting this with the variance-filtered matrix and removing the CpGs identified in the paired AH–tumor analysis, the resulting feature set 8,999 CpGs was used for training.

We implemented the classifier using gradient boosted decision trees with the xgboost package (*69*), specifying a multi-class objective (multi:softprob). Despite the resulting high-dimensional setting with substantially more features than samples, gradient boosted decision trees are well suited for this scenario due to their inherent regularization, stochastic subsampling, and robustness to high-dimensional data. Hyperparameters were fixed as follows: learning rate (eta) 0.05, maximum tree depth 6, min_child_weight 1, subsample 0.8, colsample_bytree 0.8, evaluation metric mlogloss, and 300 boosting rounds. These parameters were chosen a priori to limit model complexity and reduce overfitting in the context of high-dimensional methylation data rather than being optimized through extensive hyperparameter tuning. Missing values were handled natively by XGBoost without imputation. Five-fold cross-validation was performed with folds stratified by the interaction of class and tissue type to preserve class balance and tissue composition, and out-of-fold predictions and probabilities were recorded for all samples.

Classifier performance was evaluated using confusion matrices and one-vs-rest ROC curves generated with the pROC package (*70*). Macro-AUC (mean across classes) and micro-AUC (pooled across all classes) were calculated. To visualize the limma-based feature selection, volcano plots and boxplots of top CpGs per class were created using ggplot2 (*58*).

A final XGBoost model was retrained on all samples with the final fixed feature set of 8999 CpGs. Feature importance from this model was exported with xgboost and used to generate z-scored heatmaps with ComplexHeatmap (*53, 54*) and circlize, as well as t-SNE embeddings with Rtsne (*56*) (perplexity set heuristically as max(5, min(30, floor((n–1)/3)))), visualized by cluster and tissue type.

For independent validation, the final model was applied to the Liu cohort (*9*). As shown by Ryl et al. (*10*), subtype 1 corresponds to Cluster A, subtype 2 without *MYCN* amplification corresponds to Cluster B, and subtype 2 with *MYCN* amplification corresponds to Cluster C. Accordingly, we assigned samples from the Liu cohort (*9*) to these clusters, yielding a total of 63 samples for independent validation, consisting of 27 Cluster A, 32 Cluster B, and 4 Cluster C cases. Validation β-matrices were reordered to match the training feature order, with missing CpGs retained as NA (supported by XGBoost). Predictions were generated as class probabilities and assigned labels. Because the validation dataset did not include all training classes, confusion matrices were reported only for classes present.

Calibration of predicted class probabilities was assessed on the independent validation cohort. Probabilistic performance was summarized using the multiclass Brier score, and class-wise one-versus-rest Brier scores were calculated to evaluate the precision of probability estimates for individual retinoblastoma clusters. In addition, reliability diagrams were generated for each cluster by binning predicted class probabilities into deciles based on quantiles and comparing the mean predicted probability per bin to the observed class frequency.

### Statistical Analysis

Statistical analyses were performed in R (version 4.3.3) and using GraphPad Prism (version 10.1.0). Pearson’s correlation was used to assess associations between tumor fraction and AH liquid biopsy volume or cfDNA concentration, as well as to describe concordance between tumor tissue and AH-derived CNV profiles. Group-wise differences in AH volume, cfDNA concentration, tumor fraction and median cfDNA fragment length across clinical and pathological variables were assessed using one-way ANOVA. P values are reported without adjustment for multiple testing.

## Data Availability

All data produced in the present study are available upon reasonable request to the authors

## List of Supplementary Materials

**Figures S1 to S4**

Figure S1 Aqueous humor cfDNA reflects tumor biology through copy number, fragmentation, and mutation signatures.

Figure S2 Cluster-specific methylation patterns are preserved in aqueous humor cfDNA.

Figure S3 Integrative CNV and promoter methylation analysis across methods and clinical subgroups in retinoblastoma.

Figure S4 Machine learning-based classification of retinoblastoma subtypes using tumor and liquid biopsy methylation profiles.

**Tables S1 to S10**

Table S1 Clinical and histopathological characteristics of the retinoblastoma cohort.

Table S2 cfDNA yield, purity, and ctDNA fraction in AH samples.

Table S3 SNVs detected in AH cfDNA and RB1 variants in matched tumor tissue.

Table S4 Cross-platform harmonization and correlation of CNV profiles.

Table S5 CpG sites excluded due to biomaterial-specific methylation differences.

Table S6 Class-discriminative CpG sites identified by one-versus-rest analysis.

Table S7 Final CpG feature set used for classifier training.

Table S8 Cross-validation predictions of the XGBoost classifier.

Table S9 Independent validation predictions of the classifier using the Liu et al. cohort.

Table S10 Feature importance ranking of CpG sites in the final XGBoost classifier.

## Acknowledgments

The authors thank the High Throughput Sequencing unit of the Genomics and Proteomics Core Facility German Cancer Research Center (DKFZ), for providing excellent sequencing services and the Microarray Core Facility (DKFZ) for providing the Illumina methylation arrays and related services. The authors thank the Omics IT and Data Management Core Facility (ODCF), DKFZ, for the data management infrastructure.

## Author contributions

Conceptualization: PK, KKM, SV, SHM, TR

Methodology: PK, TR, KKM, SV, SHM

Investigation: SHM, TW, NS, MM, DH, ST, FS

Bioinformatics and statistics: SV, SHM

Sample and clinical data collection: EB, LJ, TK, NB, PR

Contributing discussions about data and findings: PK, KKM, EA, JM, MS, ISD, RJA

Visualization: SHM, SV

Supervision: PK, KKM

Funding acquisition: PK, TR

Writing - original draft: SHM, SV

Writing – review & editing: SV, SHM, PK, KKM, TR, EA, DH, JM, PP, MS, EB, MB, SMP, KWP

## Data and materials availability

The high-throughput sequencing datasets generated for this study will be made available in EGA European Genome-Phenome Archive upon acceptance of the manuscript. DNA methylation array data will be deposited in the Gene Expression Omnibus (GEO) repository upon acceptance of the manuscript. Custom code used for data processing and analysis is available from the corresponding authors upon reasonable request.

