## Supplementary material for "Aqueous Humor Liquid Biopsy Enables Multi-Omics Tumor Profiling and Methylation-Based Machine-Learning Stratification of Retinoblastoma": Figure S

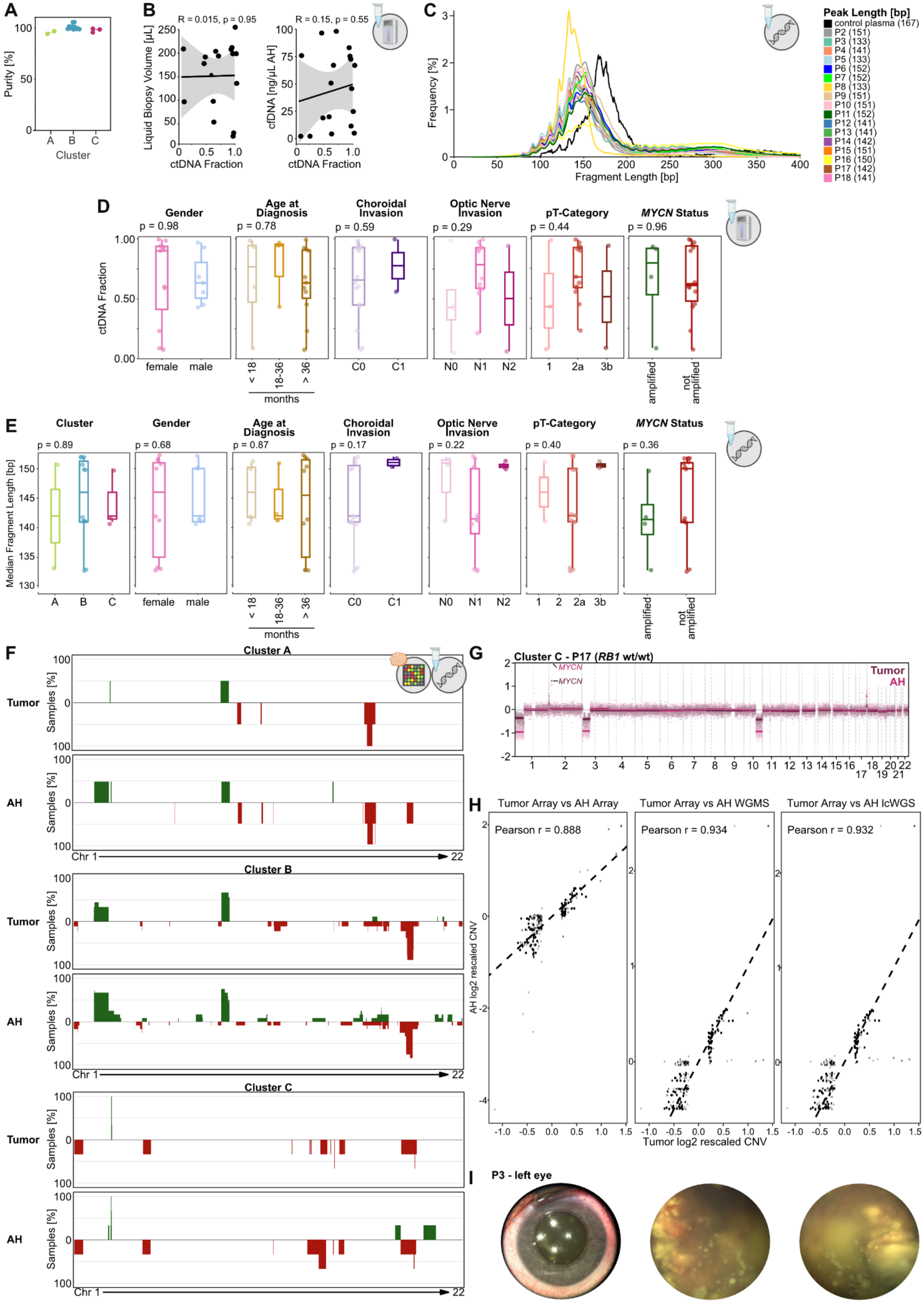
A
Purity [%]
Cluster
A B C
B
R = 0.015, p = 0.95
Liquid Biopsy Volume [µL]
ctDNA Fraction
R = 0.15, p = 0.55
cfDNA [ng/µL AH]
ctDNA Fraction
C
Frequency [%]
Fragment Length [bp]
Peak Length [bp]
control plasma (167)
P2 (151)
P3 (133)
P4 (141)
P5 (133)
P6 (152)
P7 (152)
P8 (133)
P9 (151)
P10 (151)
P11 (152)
P12 (141)
P13 (141)
P14 (142)
P15 (151)
P16 (150)
P17 (142)
P18 (141)
D
Gender
p = 0.98
Age at Diagnosis
p = 0.78
Choroidal Invasion
p = 0.59
Optic Nerve Invasion
p = 0.29
pT-Category
p = 0.44
MYCN Status
p = 0.96
ctDNA Fraction
female male
< 18 18-36 > 36
months
C0 C1
N0 N1 N2
1 2a 3b
amplified
not amplified
E
Cluster
p = 0.89
Gender
p = 0.68
Age at Diagnosis
p = 0.87
Choroidal Invasion
p = 0.17
Optic Nerve Invasion
p = 0.22
pT-Category
p = 0.40
MYCN Status
p = 0.36
Median Fragment Length [bp]
A B C
female male
< 18 18-36 > 36
months
C0 C1
N0 N1 N2
1 2 2a 3b
amplified
not amplified
F
Cluster A
Tumor
AH
Samples [%]
Chr 1
22
Cluster B
Tumor
AH
Cluster C
Tumor
AH
G
Cluster C - P17 (RB1 wt/wt)
MYCN
MYCN
Tumor
AH
H
Tumor Array vs AH Array
Tumor Array vs AH WGMS
Tumor Array vs AH lcWGS
Pearson r = 0.888
Pearson r = 0.934
Pearson r = 0.932
AH log2 rescaled CNV
Tumor log2 rescaled CNV
I
P3 - left eye

**Figure S1 Aqueous humor cfDNA reflects tumor biology through copy number, fragmentation, and mutation signatures. (A)** Estimated purity of cfDNA extracted from AH samples, stratified by retinoblastoma cluster assignment. Each dot represents one sample. **(B)** Correlation analyses between tumor fraction derived from WGMS and (left) AH liquid biopsy volume and (right) cfDNA concentration. Each dot represents one patient. Pearson correlation coefficient (R) and p-values are indicated. **(C)** Fragment length distribution of individual AH samples and control plasma. Boxplots showing **(D)** tumor fraction (from WGMS) and **(E)** median cfDNA fragment length (from lcWGS) in AH samples, stratified by clinical and pathological parameters: retinoblastoma cluster, gender, age at diagnosis, laterality, enucleation type, choroidal invasion, optic nerve invasion, pT category, and MYCN amplification status. P-values were calculated using one-way ANOVA. **(F)** Fusion plots of copy number alterations per chromosome for Cluster A, B and C, shown separately for tumor and AH. The y-axis indicates the percentage of samples with gains (green) or losses (red) at each genomic locus. Tumor CNVs were derived from methylation array data; AH CNVs from lcWGS. **(G)** CNV plot of of patient 17 (P17), overlayed for tumor (dark) and AH (light) samples. **(H)** Pooled scatterplots showing Pearson correlation between rescaled CNV profiles derived from AH cfDNA and matched tumor tissue across all informative autosomal bins (absolute tumor $\log_2$ copy-number ratio > 0.2), shown separately for array-based AH, WGMS-AH, and lcWGS-AH. Each point represents one patient–bin pair. **(I)** RetCam® pictures of the left eye of patient 3 (P3).

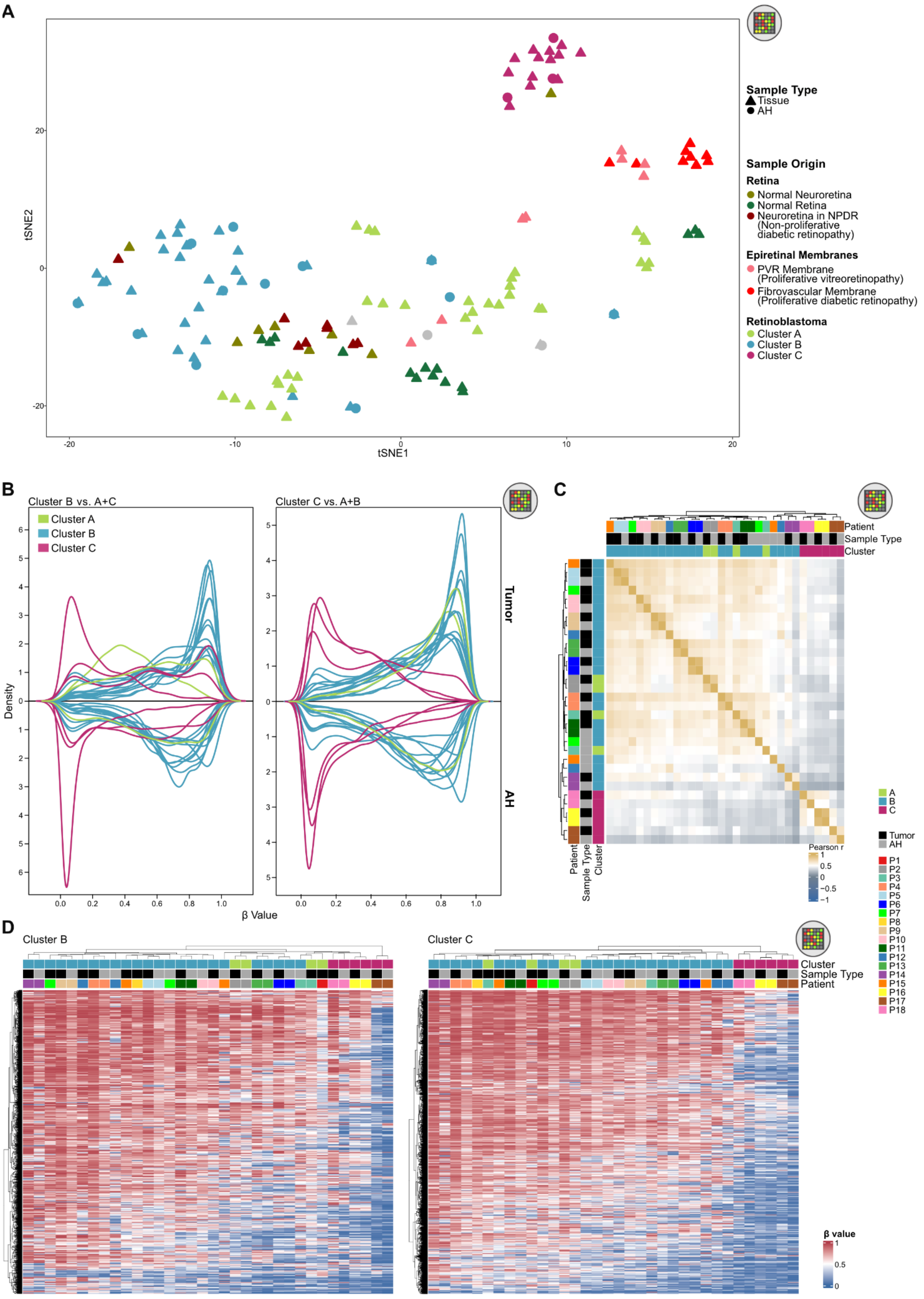
A
Sample Type
Tissue
AH
Sample Origin
Retina
Normal Neuroretina
Normal Retina
Neuroretina in NPDR
(Non-proliferative diabetic retinopathy)
Epiretinal Membranes
PVR Membrane
(Proliferative vitreoretinopathy)
Fibrovascular Membrane
(Proliferative diabetic retinopathy)
Retinoblastoma
Cluster A
Cluster B
Cluster C
tSNE1
tSNE2
B
Cluster B vs. A+C
Cluster C vs. A+B
Cluster A
Cluster B
Cluster C
Density
β Value
Tumor
AH
C
Patient
Sample Type
Cluster
Pearson r
A
B
C
Tumor
AH
P1
P2
P3
P4
P5
P6
P7
P8
P9
P10
P11
P12
P13
P14
P15
P16
P17
P18
D
Cluster B
Cluster C
Cluster
Sample Type
Patient
β value

**Figure S2 Cluster-specific methylation patterns are preserved in aqueous humor cfDNA. (A)** t-SNE plot of tumor and AH samples based on cluster-discriminating CpG sites identified by Ryl et al. The dataset includes additional tumor and control samples from Ryl et al. (10), Liu et al. (9), and Berdasco et al. (49). Points are color-coded by sample origin or classification and shaped by tissue type (tissue vs. AH). **(B)** Density plots of β-values across cluster-discriminating CpG sites identified by Ryl et al., shown for individual AH samples. (Left), CpGs separating Cluster B from Clusters A and C. (Right), CpGs separating Cluster C from Clusters A and B. Each line represents one AH sample; lines are color-coded by retinoblastoma cluster. CpG sites are the same as in Figure 3B. **(C)** Heatmap of pairwise Pearson correlations among tumor and AH samples, based on cluster-discriminating CpG sites identified by Ryl et al. All 30 samples (15 tumor, 15 AH) were correlated with one another and hierarchically clustered. Annotations indicate patient identity, sample type, and cluster assignment. **(D)** Heatmaps of β-values for CpG sites discriminative for (left) Cluster B versus Clusters A and C, and (right) Cluster C versus Clusters A and B, as identified by Ryl et al. Rows represent CpG sites; columns represent AH and tumor samples derived from methylation array data. Samples are annotated by patient, sample type, and retinoblastoma cluster assignment.

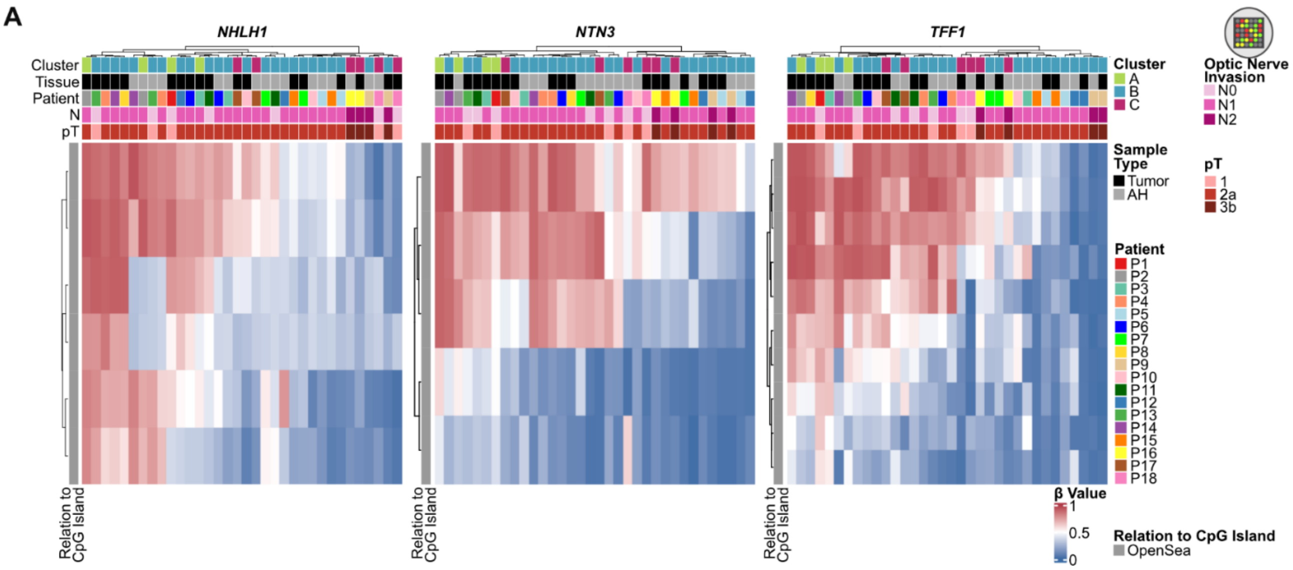


**Figure S3 Integrative CNV and promoter methylation analysis across methods and clinical subgroups in retinoblastoma. (A)** Heatmaps of β-values for CpG sites within the promoter regions *of NHLH1*, *NTN3*, *and TFF1*, based on methylation array data. Rows represent CpG sites; columns represent tumor and AH samples. Samples are annotated by patient, retinoblastoma cluster, sample type, optic nerve invasion, and pT category.

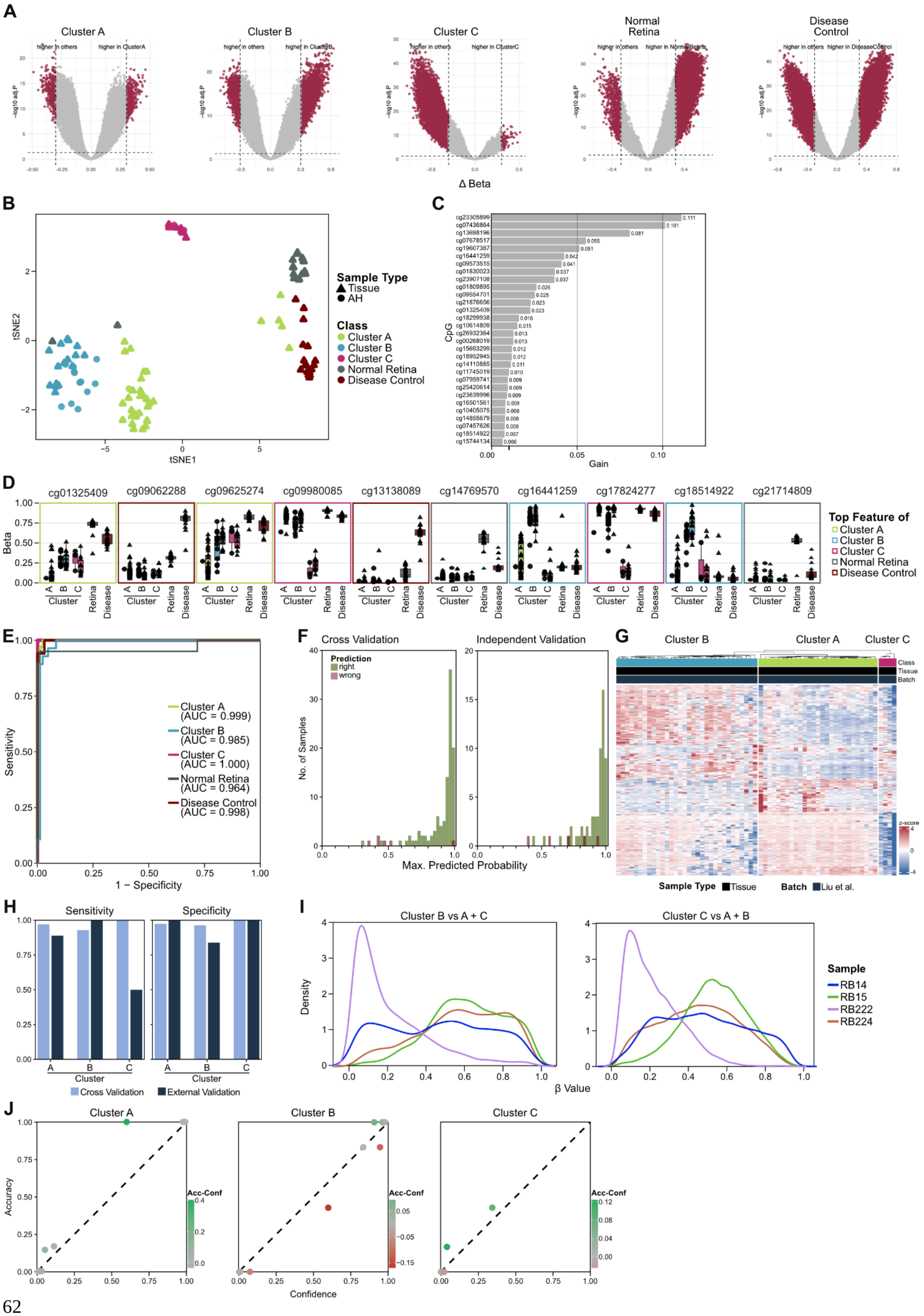
A
Cluster A
Cluster B
Cluster C
Normal Retina
Disease Control
−log10 adj.P
Δ Beta
B
tSNE1
tSNE2
Sample Type
Tissue
AH
Class
Cluster A
Cluster B
Cluster C
Normal Retina
Disease Control
C
CpG
Gain
D
cg01325409
cg09062288
cg09625274
cg09980085
cg13138089
cg14769570
cg16441259
cg17824277
cg18514922
cg21714809
Beta
Cluster
Retina
Disease
Top Feature of
Cluster A
Cluster B
Cluster C
Normal Retina
Disease Control
E
Sensitivity
1 − Specificity
Cluster A (AUC = 0.999)
Cluster B (AUC = 0.985)
Cluster C (AUC = 1.000)
Normal Retina (AUC = 0.964)
Disease Control (AUC = 0.998)
F
Cross Validation
Independent Validation
Prediction
right
wrong
No. of Samples
Max. Predicted Probability
G
Cluster B
Cluster A
Cluster C
Class
Tissue
Batch
z-score
Sample Type
Tissue
Batch
Liu et al.
H
Sensitivity
Specificity
Cluster
Cross Validation
External Validation
I
Cluster B vs A + C
Cluster C vs A + B
Density
β Value
Sample
RB14
RB15
RB222
RB224
J
Cluster A
Cluster B
Cluster C
Accuracy
Confidence
Acc-Conf

**Figure S4 Machine learning-based classification of retinoblastoma subtypes using tumor and liquid biopsy methylation profiles. (A)** Volcano plots showing differentially methylated CpGs of each class identified by one versus rest limma analysis for each class. **(B)** Informed t-SNE clustering based on the 300 most important features. Sample type is annotated by shape, class by color. **(C)** Barplot showing the top 30 CpG sites ranked by mean importance in the random forest classifier trained on methylation array data. **(D)** Boxplots of β-values for the two most important CpG sites per class identified by the XGBoost model, color-coded by class. **(E)** ROC curves for the classifier, using a one-vs-rest approach for each class (Cluster A, B, C, normal retina, and disease control). Curves are color-coded by class, and AUC values are shown in the legend. **(F)** Maximum predicted probability per sample for (left) cross-validation and (right) external validation. Correctness of prediction is shown by color (green=correct, red=wrong). **(G)** Correlation heatmap of the top 300 most important CpGs from the final model. Rows represent CpG sites; columns represent tissue and AH samples. Samples are annotated by class, sample type and batch. **(H)** Comparison of sensitivity and specificity of the classifier detecting the individual classes in cross validation and external validation. **(I)** Density plots of β-values across cluster-discriminating CpG sites identified by Ryl et al., shown for all MYCN-amplified samples in the Liu et al. cohort. (Left), CpGs separating Cluster B from Clusters A and C. (Right), CpGs separating Cluster C from Clusters A and B. Lines are color-coded by sample. **(J)** Calibration analysis showing accuracy over confidence for Cluster A, B and C.
